# Towards a participatory European social contact observatory

**DOI:** 10.1101/2025.11.17.25338859

**Authors:** Kathleen N. Kelley, Jantien Backer, Pietro Coletti, Nicolò Gozzi, Lisa Hermans, Niel Hens, Neilshan Loedy, Mattia Mazzoli, Andrea Torneri, Albert Jan van Hoek, Daniela Paolotti

## Abstract

Social contacts shape infectious disease transmission, but most contact data come from infrequent, resource-intensive stand-alone surveys that are difficult to link with longitudinal health information. Digital participatory surveillance offers an alternative for continuous collection of data requiring fewer resources. We analyzed over 110,000 contact reports from 18,000 InfluenzaNet participants, linked with sociodemographic, behavioral, and symptom data. Weighted adult age-stratified contact matrices reproduced established structural features, while temporal analyses showed that transmission-relevant patterns varied over time. Negative binomial mixed-effects models found fewer contacts among participants with influenza-like illness, particularly ILI with fever, although milder symptoms were associated with more contacts in one Dutch specification. Contacts also varied with employment, weekends, and holidays. Participatory surveillance can therefore generate valid, longitudinal contact data while integrating otherwise difficult-to-obtain participant information. This infrastructure provides a foundation for a European social contact observatory capable of providing regularly updated inputs for epidemic modeling and public health preparedness.

## 1 Introduction

Understanding how people interact is central to modeling and predicting the spread of infectious diseases ^1^. Social contacts, typically defined as close-range face-to-face encounters, provide opportunities for transmission via droplets and aerosols, key pathways for respiratory pathogens such as influenza and SARS-CoV-2. Integrating realistic contact patterns into epidemic models of these types of infections improves their accuracy and interpretability ^2–8^. By monitoring variations in contact volume and structure, this data is crucial for capturing behavioral changes linked to disease spread ^9^ and evaluating the impact of non-pharmaceutical interventions (e.g., social distancing) on transmission dynamics ^10–14^. More recently, contact data has been used to assess how socio-economic factors shape disparities in disease burden by examining differences in exposure across groups ^15–17^, such as occupational groups that vary in their ability to reduce contacts by working from home.

Social contacts can be measured in several ways, including proximity sensors ^18–21^ and time use surveys ^22–24^, or inferred from large-scale synthetic models ^25;26^. Nonetheless, contact diary surveys remain the primary tool used for collecting data on social interactions ^27^. In these studies, participants report their encounters on a given day, recording attributes such as (perceived) age and gender of each contacted individual, and setting for each contact (i.e., workplace, school, etc). The main output of these studies is the social contact matrix, i.e. a matrix detailing the average number of contacts between individuals (usually on a daily basis) depending on their age. The landmark POLYMOD study (2006) collected this information via a paper-based survey across eight European countries ^28^, providing age-structured contact matrices that have since been widely used in transmission models of various infectious diseases ^29–31^.

Almost fifteen years later, during the COVID-19 pandemic, contact patterns from studies conducted under stable public health conditions, such as POLYMOD, no longer reflected the behavior of individuals living through a public health crisis marked by widespread restrictions. The introduction of non-pharmaceutical interventions aimed at curbing the spread of SARS-CoV-2 significantly impacted both the overall number and the structure of social contacts. For this reason, several novel data collections were rapidly deployed to capture contact patterns under evolving public health landscapes and social distancing measures ^9;32;33^. A notable example is the CoMix digital survey launched in March 2020 that collected social contact data over multiple waves for over 20 European countries between 2020 and 2021^34^, with additional rounds later conducted in several countries in 2022^35^.

While highly informative and valuable, traditional, stand-alone contact surveys face significant challenges: they are logistically demanding, costly, and resource-intensive, as they require extensive recruitment, training, and data collection. As a result, they are typically conducted infrequently and with a cross-sectional approach, limiting their ability to monitor contact behavior across time and changing public health environments. Additionally, due to the level of required detail in the contact-related questions (i.e. participants must record every encounter), surveys often restrict the scope and duration of data collection to reduce participant burden. This limits the opportunity to capture richer information about underexplored determinants of contacts and longitudinal patterns that could provide deeper insights into the drivers of contact behavior and enable extended representations of contact matrices beyond the commonly considered dimension of age ^36^.

Digital participatory surveillance platforms, by leveraging the voluntary participation of citizens, help address the challenges faced by traditional contact surveys. Indeed, they offer direct access to a pool of individuals that can be routinely probed to capture health and behavioral data in a cost-effective manner. InfluenzaNet, present in 12 European countries, is a consortium of digital participatory surveillance platforms that has continuously collected data through voluntary, web-based participation since 2009^37;38^. Over the years, the InfluenzaNet platforms have become reliable sources of high-quality, longitudinal data on influenza-like illness (ILI) prevalence, vaccination rates, and health-related behaviors, among other indicators ^39–42^.

In this work, we aim to test the feasibility of using participatory surveillance platforms to collect contact data for epidemiological applications. In this setting, comparison with established contact studies is useful for assessing whether the platform captures plausible age-mixing patterns, whereas repeated measurements over time are needed to demonstrate its value as a surveillance tool. To this aim, a digital contact survey was developed and administered to the users of three InfluenzaNet ^40^ platforms: Influweb in Italy and the Infectieradar platforms operating in Belgium and the Netherlands. Users were invited to submit a contact diary every two months, reporting the number of contacts during the previous day with individuals in different age groups in four settings. By building on the existing infrastructure, this required minimal additional logistics, recruitment effort, and substantially fewer resources than a new stand-alone survey. The platform’s integration of concurrent surveys allowed us to link participants’ contact responses with their questionnaires on socio-demographics, self-reported symptoms, and health-related behaviors, providing a more comprehensive picture of health and behavior while only minimally increasing respondent burden. Moreover, the platform’s capacity for continual data collection means that longitudinal trends and shifts in contact patterns during periods of heightened disease awareness or altered public health conditions, well documented during COVID-19 ^33;43^ and earlier pandemics ^44^, can be monitored as they occur, providing insights not possible with one-off surveys.

We enriched over 110, 000 contact reports from more than 18, 000 unique users across three European countries, with information regarding self-reported symptoms, socio-demographics, and health-related behaviors of each participant. We constructed overall and setting-specific age-stratified contact matrices and compared them with prior established studies, finding broadly similar age-mixing patterns, including strong age-assortative mixing. We then modeled individual-level drivers of the reported number of contacts, recovering associations previously observed in other diary-based contact studies, while incorporating participant health reports to assess symptom-related variation in contact behavior. Our work demonstrates the potential of digital participatory surveillance for the longitudinal monitoring of social contacts and illustrates its unique added value in linking contact behavior with self-reported symptoms and health-related behaviors.

## 2 Materials and Methods

### 2.1 Ethics statement

Ethical approval and informed consent procedures for the participating platforms were conducted in accordance with the national regulations applicable in each country. Participation in all InfluenzaNet platforms was voluntary, and electronic informed consent was obtained from participants at registration prior to data collection. Participants were informed about the purpose of the surveillance systems, the processing and storage of personal data, and their right to withdraw from participation at any time. Data is anonymized prior to analysis and handled in accordance with applicable national legislation and the General Data Protection Regulation (GDPR).

### 2.2 Study design and data

Influenzanet platforms

InfluenzaNet is a network of European participatory digital surveillance platforms for monitoring influenza-like illness (ILI) ^45^, of which Infectieradar (Belgium), Infectieradar (the Netherlands), and Influweb (Italy) form part. This participatory surveillance network operates in parallel with traditional public health surveillance systems, with platforms active in Belgium and the Netherlands since 2003 and in Italy since 2008. The signal from InfluenzaNet’s syndromic surveillance has been shown to align well with those from sentinel-based systems, supporting its utility as a data source ^37;38;45^.

The data used in this study are based on three types of recurring surveys hosted on the platforms:

1. Intake survey: This collects demographic and health information, including age, sex, region of residence, education level, vaccination status, and the presence of chronic conditions. Participants are invited every year to submit new intake information.
2. Weekly syndromic survey: Participants receive weekly email reminders to report whether they have experienced any symptoms in the past seven days and to describe associated health behaviors. These surveys are available year-round to support continuous monitoring of ILI.
3. Social contact survey: Beginning in October 2023 (the Netherlands), February 2024 (Italy), and October 2024 (Belgium) participants enrolled in the syndromic survey were invited to take part in a social contact study. The survey prompts individuals to report all interpersonal contacts that occurred between 5:00 a.m. the previous day and 5:00 a.m. on the day of survey completion. Reported contacts are categorized by age group, gender of the contact, and the setting in which the contact took place (home, work, school, leisure).

This is an observational longitudinal study, with data collection ongoing at the time of writing. Participants were invited to complete a contact survey every other month, while continuing to receive invitations to fill in weekly syndromic surveillance surveys. This bi-monthly schedule was intended to minimize participant burden and reduce the likelihood of survey fatigue. Recruitment was rolling, meaning participants began at different times, so follow-up points do not align across individuals. For example, someone who submitted their first contact survey in March would be re-invited in May, while a participant beginning in February would receive their next invitation in April.

Comparison studies

To contextualize the findings from the InfluenzaNet contact surveys, we compared our results with data from representative prior studies of social contact behavior. Two widely used, multi-country datasets are POLYMOD ^28^, a pre-pandemic European contact survey, and CoMix ^34^, a longitudinal contact survey conducted during and after the COVID-19 pandemic. In both cases, we used only the data collected in either Belgium, Italy or the Netherlands.

Country-specific comparator studies were also included where available: the Flemish Contact Survey and EPICURUS for Belgium ^46;47^, MixIT for Italy ^10^, and the Pienter2, Pienter3, and PienterCorona contact studies for the Netherlands ^48;49^.

Further details on study design, sample sizes, timing, and inclusion decisions are provided in Table 1.

**Table 1.** Comparator social contact studies included in the analysis. Summary of comparator datasets used to contextualize contact matrices from the InfluenzaNet platforms. Data publically available at socialcontactdata.org.

| Study | Country | Timing | Sample | Data collection | Study design and inclusion notes |
| --- | --- | --- | --- | --- | --- |
| POLYMOD<sup>28;50</sup> | Belgium | 2005–2006 | 750 | Paper | Cross-sectional diary survey. |
|  | Italy | 2005–2006 | 849 | Paper | Cross-sectional diary survey. |
|  | Netherlands | 2005–2006 | 269 | Paper | Cross-sectional diary survey. |
| CoMix<sup>34;35;51;52</sup> | Belgium | 2022 | 1500 | Online | Only post-pandemic contact survey round used. |
|  | Italy | 2021 | 7657 obs./2004 part. | Online | Longitudinal survey conducted during active COVID-19 public health interventions. |
|  | Netherlands | 2022 | 1491 | Online | Only post-pandemic contact survey round used. |
| Flemish Contact Survey<sup>46;53</sup> | Belgium | 2010–2011 | 1752 | Paper | Survey of the Flemish region of Belgium. |
| EPICURUS<sup>47;54</sup> | Belgium | 2022–2023 | 5723 | Multi-modal: paper, online, in-person interviews | Sample including the general population and selected target populations: residents of care facilities, individuals with chronic illnesses, and individuals with ILI. |
| MixIT<sup>10;55</sup> | Italy | 2022–2023 | 3743 | Online | Only direct contacts, not indirect, were included in this study. |
| Pienter2<sup>48</sup> | Netherlands | 2006–2007 | 8465 | Paper | Seroepidemiological survey with social contact data. |
| Pienter3<sup>49;56</sup> | Netherlands | 2016–2017 | 5574 | Paper | Seroepidemiological survey with social contact data. |
| PienterCorona<sup>49;56</sup> | Netherlands | 2022–2023 | 21811 | Online | Longitudinal study; only data collected during 2022 and 2023 were included. |

### 2.3 Age-based sample restriction

For participants younger than 19 years, survey responses were provided by a parent or guardian. Given the small number of minor participants relative to the total sample size within each country, analyses were restricted to adult participants in order to avoid unstable estimates based on sparse cell counts. The age distribution of the sample is presented in Table 2 of Section 3.1. Sensitivity analyses assessing the impact of this restriction are presented in the Supporting Information.

**Table 2.** Demographic overview of study participants and survey responses. Descriptive statistics for study participants and survey responses from the Infectieradar platform in Belgium, Influweb in Italy, and the Infectieradar platform in the Netherlands. Participant levels marked with *^∗^* were excluded from regression models and contact matrix estimation. Note percentages may not sum to 100% due to rounding.

|  | No. of participants (%) |  |  | No. of responses (%) |  |  |
| --- | --- | --- | --- | --- | --- | --- |
|  | BE | IT | NL | BE | IT | NL |
| Age group |  |  |  |  |  |  |
| 0–19 | 6 (1%)* | 23 (5%)* | 106 (1%)* | 23 (1%)* | 96 (4%)* | 409 (< 1%)* |
| 20–49 | 148 (19%) | 143 (29%) | 2902 (22%) | 392 (17%) | 607 (23%) | 15951 (17%) |
| 50–69 | 393 (50%) | 249 (51%) | 7674 (58%) | 1159 (50%) | 1417 (54%) | 54170 (58%) |
| 70+ | 246 (31%) | 78 (16%) | 2600 (20%) | 749 (32%) | 483 (19%) | 23004 (25%) |
| Sex |  |  |  |  |  |  |
| Female | 459 (58%) | 222 (45%) | 7467 (56%) | 1329 (57%) | 1103 (42%) | 51520 (55%) |
| Male | 334 (42%) | 271 (55%) | 5815 (44%) | 994 (43%) | 1500 (58%) | 42014 (45%) |
| Education |  |  |  |  |  |  |
| High school or less | 197 (25%) | 246 (50%) | 4893 (37%) | 607 (26%) | 1269 (49%) | 33239 (36%) |
| University | 596 (75%) | 247 (50%) | 8389 (63%) | 1716 (74%) | 1334 (51%) | 60295 (64%) |
| Employment status |  |  |  |  |  |  |
| Student | 9 (1%)* | 44 (9%) | 191 (1%)* | 29 (1%)* | 198 (8%) | 840 (1%)* |
| Employed | 319 (40%) | 281 (57%) | 7545 (57%) | 886 (38%) | 1412 (54%) | 51093 (55%) |
| Unemployed | 62 (8%) | 33 (7%) | 1384 (10%) | 174 (7%) | 187 (7%) | 9572 (10%) |
| Retired | 403 (51%) | 135 (27%) | 4162 (31%) | 1234 (53%) | 806 (31%) | 32029 (34%) |
| Household size |  |  |  |  |  |  |
| 0 | 33 (4%) | 23 (5%) | 2231 (17%) | 88 (4%) | 100 (4%) | 16204 (17%) |
| 1–3 | 649 (82%) | 348 (71%) | 10434 (79%) | 1945 (84%) | 1918 (74%) | 73791 (79%) |
| 4+ | 111 (14%) | 122 (25%) | 617 (5%) | 290 (12%) | 585 (22%) | 3539 (4%) |
| Chronic condition |  |  |  |  |  |  |
| No | 556 (70%) | 353 (72%) | 9685 (73%) | 1625 (70%) | 1776 (68%) | 66576 (71%) |
| Yes | 237 (30%) | 140 (28%) | 3597 (27%) | 698 (30%) | 827 (32%) | 26958 (29%) |
| Holiday period |  |  |  |  |  |  |
| Non-holiday | — | — | — | 1664 (72%) | 1779 (68%) | 64296 (69%) |
| Holiday | — | — | — | 659 (28%) | 824 (32%) | 29238 (31%) |
| Survey day |  |  |  |  |  |  |
| Weekend | — | — | — | 636 (27%) | 805 (31%) | 30064 (32%) |
| Weekday | — | — | — | 1687 (73%) | 1798 (69%) | 63470 (68%) |

### 2.4 Post-stratification weights

Post-stratification weights were applied to align the study samples with the corresponding reference populations, defined as the Belgian, Italian, or Dutch general population in the year of data collection. Weights accounted for differences in the age and sex distribution of the survey sample relative to the reference population, as well as differences in the distribution of responses between weekdays and weekends.

The weights were applied to the InfluenzaNet samples in all analyses, including contact matrix construction, regression modeling, and estimation of median daily contacts by participant characteristics. The same weighting approach was also applied when constructing contact matrices from comparator datasets (e.g., POLYMOD, CoMix) to ensure consistency and comparability across studies.

Further details on the calculation of post-stratification weights are provided in the Supplementary Information.

### 2.5 Contact matrix construction

Weighted contact rates were computed as the mean number of contacts between respondent and contact age groups. The following age groups were defined for use in the contact matrices: 19–29, 30–39, 40–49, 50–59, 60–69, and 70 years and older. For respondent age group *i* and contact age group *j*:

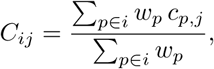

where *c_p,j_*is the number of contacts reported by respondent *p* with individuals in group *j*, and *w_p_* is the participant weight, described in section 2.4.

As social interactions are reciprocal, we symmetrize matrices using census-based estimates of the country’s population in the survey year ^57–59^. Let *P_i_* (*P_j_*) denote the population size of age group *i* (*j*). The symmetric contact rate is ^17;60^:

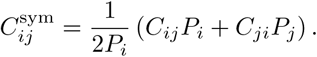

The exclusion of participants younger than 19 years from the analytic sample means that reciprocity can only be imposed among adult respondent age groups, while contacts reported by adult respondents with individuals younger than 19 years were retained as an additional contact age-group column. The resulting matrices are rectangular, consisting of a symmetrized adult-by-adult component and a non-symmetrized column of contacts from adult respondents to minors.

Uncertainty was estimated using bootstrap resampling. For each study, participants were sampled with replacement and contact matrices were reconstructed using the procedure above. Final matrices were calculated as the element-wise mean of the bootstrap replicates, with confidence intervals estimated from the bootstrap distribution ^61^. All procedures were implemented using the socialmixr R package ^62;63^.

### 2.6 Modeling number of contacts

#### 2.6.1 Variable description

The modeling analysis included a set of variables commonly identified in the literature as associated with contact behavior ^16;28;64;65^. Age was categorized into groups: 19–49, 50-69 (reference category), and 70 years and older. Sex was coded as a binary variable (male or female), and employment status was classified into levels: employed (reference), student, unemployed, and retired. The day of the week on which contacts were reported was included as a binary indicator for weekday. Household size reflects the number of people living in the respondent’s home and educational attainment was included as a categorical variable with two levels: high school or less and university education.

Additionally, we included an indicator for whether the respondent report taking medication for a chronic condition, such as asthma, diabetes, cardiovascular or pulmonary disease, renal disease, or conditions involving immunosuppression. To account for potential temporal variation in contact behavior, we included an indicator variable for whether the contact survey was submitted during a school or public holiday period. Holiday periods were defined based on national public holidays and academic calendars.

Finally, we made use of the rich data we have on all contact survey participants by matching each contact survey to a corresponding weekly symptom report, a process described in section 2.6.2. Using the ECDC case definition ^66^, we classified symptom history in the 30 days preceding the contact survey into four categories: no symptoms (reference), symptoms not qualifying as ILI, symptoms qualifying as ILI, and symptoms qualifying as ILI including fever.

#### 2.6.2 Data preparation

To incorporate symptom-related variables into the analysis of contact patterns, we merged the contact survey dataset with the weekly syndromic surveillance dataset. Symptom-based analyses were restricted to contact survey entries from participants who had submitted at least one symptom survey in the 30 days before the contact survey, to avoid treating the absence of a recent symptom report as evidence of no symptoms. This restriction did not substantially reduce the sample size in any country.

Each contact survey entry was linked to symptom reports from the same participant. As participants could submit multiple symptom surveys within the 30-day window (one per week), we assigned symptom status to each contact survey entry using two approaches:

- *Temporal proximity*: selecting the symptom survey submitted closest in time prior to the contact survey, which often occurred on the same day (see Table 3).
- *Symptom burden*: selecting the symptom survey with the highest number of reported symptoms within the prior 30 days.

**Table 3.** Overview of symptoms among survey responses. Descriptive statistics for responses from the Infectieradar platform in Belgium, Influweb in Italy, and the Infectieradar platform in the Netherlands. Weekly symptom reports in the 30 days prior to contact diary reports were matched using either the temporal proximity approach or the symptom burden approach.

|  | No. of responses (%) |  |  |  |  |  |
| --- | --- | --- | --- | --- | --- | --- |
|  | BE |  | IT |  | NL |  |
|  | Sym. burden | Temp. prox. | Sym. burden | Temp. prox. | Sym. burden | Temp. prox. |
| *Prioritizing:* |  |  |  |  |  |  |
| Symptom count |  |  |  |  |  |  |
| 0 | 1400 (62%) | 1833 (81%) | 1918 (74%) | 2308 (89%) | 60245 (65%) | 76370 (82%) |
| 1 | 221 (10%) | 104 (5%) | 177 (7%) | 75 (3%) | 8250 (9%) | 5322 (6%) |
| 2–4 | 422 (19%) | 218 (10%) | 322 (12%) | 145 (6%) | 15901 (17%) | 7839 (8%) |
| 5–8 | 157 (7%) | 69 (3%) | 134 (5%) | 45 (2%) | 6671 (7%) | 2779 (3%) |
| 9+ | 51 (2%) | 27 (1%) | 44 (2%) | 22 (1%) | 2118 (2%) | 875 (1%) |
| ILI symptom severity |  |  |  |  |  |  |
| Symptomatic, no ILI | 642 (29%) | 322 (14%) | 489 (19%) | 217 (8%) | 22100 (24%) | 11853 (13%) |
| ILI | 165 (7%) | 79 (4%) | 93 (4%) | 38 (1%) | 7786 (8%) | 3890 (4%) |
| ILI with fever | 44 (2%) | 17 (1%) | 95 (4%) | 32 (1%) | 3054 (3%) | 1072 (1%) |
| Days since symptom report |  |  |  |  |  |  |
| 0 | 1710 (76%) | 2223 (99%) | 2097 (81%) | 2550 (98%) | 65246 (70%) | 84588 (91%) |
| 1–7 | 198 (9%) | 20 (1%) | 125 (5%) | 28 (1%) | 7873 (8%) | 5699 (6%) |
| 8–14 | 88 (4%) | 1 (< 1%) | 127 (5%) | 11 (< 1%) | 6836 (7%) | 2103 (2%) |
| 15–21 | 130 (6%) | 7 (< 1%) | 114 (4%) | 2 (< 1%) | 5670 (6%) | 512 (1%) |
| 22–30 | 125 (6%) | 0 | 132 (5%) | 4 (< 1%) | 7560 (8%) | 283 (< 1%) |

Models were fit and compared under both matching approaches. Information from a participant’s most recent intake survey was then linked to the contact survey data.

#### 2.6.3 Negative binomial regression model

We modeled the total number of contacts reported in each country’s contact survey using negative binomial mixed-effects models implemented in glmmTMB ^67^. Participant-level random intercepts were included to account for repeated survey responses, and contact counts were truncated at 100 to reduce the influence of extreme outliers.

The model was specified as:

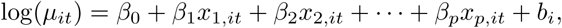

where *µ_it_*is the expected number of contacts for participant *i* on occasion *t*, *x_j,it_*are covariates, and *b_i_*is a participant-specific random intercept.

Zero inflation was assessed using likelihood ratio tests and AIC, and retained only when it improved model fit. Based on these criteria, a zero-inflated negative binomial model was used only for the Influweb Italy sample. Model estimates are reported as incidence rate ratios (IRRs), obtained by exponentiating the fitted coefficients.

### 2.7 Matrix comparison metrics

Contact matrices from the InfluenzaNet platforms were compared with matrices from the comparator social contact studies described in Table 1. These comparisons were used to assess the extent to which contact patterns estimated from the InfluenzaNet platforms were consistent with those observed in prior studies. Comparisons were made separately by country and, where available, by contact setting. Matrix similarity was evaluated using complementary metrics, each capturing a different aspect of comparison: overlap in contact volume, similarity in age-mixing pattern, and overall magnitude of difference. For the purposes of validating participatory surveillance as a data stream for epidemic modeling, similarity in relative age-mixing structure is of particular importance, whereas absolute differences in contact intensity should be interpreted in light of differences in study design, sampling frame, and social context.

#### 2.7.1 Generalized Sørensen–Dice index

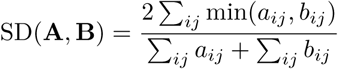

This metric quantifies overlap between two contact matrices. The index ranges from 0 (no overlap) to 1 (perfect agreement) and reflects the proportion of shared contact volume, independent of absolute scale ^68–70^.

#### 2.7.2 Cosine similarity

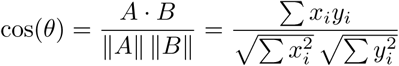

Cosine similarity measures the directional alignment between two matrices, independent of their magnitude ^71;72^. The result ranges from 0 to 1 for non-negative data, with values closer to 1 indicating that the same matrix positions tend to have relatively high or low values, or both matrices share a similar age-mixing pattern.

#### 2.7.3 Frobenius distance

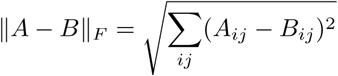

Frobenius distance captures the overall magnitude of difference between two contact matrices, producing a single non-negative number. Unlike the previous metrics, it is sensitive to the absolute scale: large differences in high-contact cells contribute more to the distance than small differences elsewhere. A value of 0 indicates identical matrices, while larger values reflect greater divergence in contact frequencies.

#### 2.7.4 Comparison plots

In a method outlined in Prem et. al 2021^26^, each contact matrix was normalized so that the scaled matrix has a dominant eigenvalue of 1. This normalization requires square matrices, so plots were constructed using the adult-by-adult matrices. After normalization, corresponding cell values from the two matrices were plotted: the normalized value from matrix A corresponds to the x-axis and the normalized value from matrix B the y-axis. A Pearson’s correlation coefficient was computed to assess agreement in cell pairs across matrices.

### 2.8 Distributions of contact intensity over time

To evaluate temporal changes in the transmission-relevant structure of social contact patterns, we estimated the distribution of spectral radii across time for each country. The spectral radius, defined as the largest absolute eigenvalue of a matrix, is directly related to the basic reproduction number, *R*_0_, in structured epidemic models and therefore provides a useful summary of transmission potential under otherwise fixed epidemiological assumptions ^73^.

Contact survey data was grouped over time using country-specific date strata. Belgium was grouped by season-year, Italy by two-month periods, and the Netherlands by month-year. These groupings were chosen to balance temporal resolution with the need for sufficient observations within each adult age class when generating stable bootstrapped contact matrices. Within each date stratum, bootstrap contact matrices were generated as described in Section 2.5, using the square, adult-by-adult component. The spectral radius was then calculated for each bootstrap matrix, producing a distribution of spectral radii over time. These distributions were plotted by date and country to assess how the transmission-relevant signal in social mixing changed over time. This analysis was intended to demonstrate the longitudinal nature of the InfluenzaNet contact survey data and to illustrate the value of ongoing data collection for capturing temporal variation in population contact patterns.

## 3 Results

### 3.1 Descriptive overview of sample and reported contacts

The study included 2,323 survey responses from 793 Infectieradar participants in Belgium, 2,603 responses from 493 Influweb participants in Italy, and 93,534 responses from 13,282 Infectieradar participants in the Netherlands. Participants were invited to complete repeated social contact surveys, with rolling recruitment resulting in survey timing that did not align across individuals. Further details on the contact survey design are provided in the Materials and Methods section 2.2.

Sample composition differed across countries. The Belgian and Dutch samples were majority female, whereas the Italian sample was majority male. University education was reported by 75% of Belgian participants, 50% of Italian participants, and 63% of Dutch participants. Participants aged 50–69 years represented the largest age group in all three samples: 50% in Belgium, 51% in Italy, and 58% in the Netherlands. Participants aged 70 years and older accounted for 31%, 16%, and 20% of the Belgian, Italian, and Dutch samples, respectively, while participants younger than 20 years represented no more than 5% of each sample. These low-count age strata were excluded from regression models and contact matrix estimation. Students were also excluded from regression models in Belgium and the Netherlands because this category was primarily composed of minor participants. Sensitivity analyses assessing the impact of these exclusions are presented in the Supporting Information.

Most survey responses were submitted on weekdays, accounting for 73% of responses in Belgium, 69% in Italy, and 68% in the Netherlands. Holiday-period responses accounted for 28%, 32%, and 31% of responses in Belgium, Italy, and the Netherlands, respectively. Further participant- and response-level characteristics are presented in Table 2.

Symptom information was linked to contact survey responses using weekly symptom reports submitted within the 30 days before the contact diary. Multiple symptom reports could be available within this window, so symptom status was summarized using two matching approaches: prioritizing the report with the highest number of reported symptoms (symptom burden) and selecting the symptom report submitted closest in time before the contact survey (temporal proximity). When symptom reports were matched using the symptom burden approach, at least one symptom was linked to 38% of Belgian responses, 26% of Italian responses, and 35% of Dutch responses. When matched using the temporal proximity approach, these proportions were lower: 19%, 11%, and 18%, respectively. Symptom distributions by country and matching approach are shown in Table 3.

On average, 155 responses were collected each month in Belgium, 105 in Italy, and 3,682 in the Netherlands. Most participants completed four or more bi-monthly contact surveys in Belgium, seven or more in Italy, and eight or more in the Netherlands during the study period, reflecting differences in the timing of study initiation across countries. Data collection began in October 2024 in Belgium, February 2024 in Italy, and October 2023 in the Netherlands.

To examine how social contact patterns vary across individual characteristics, we calculated the weighted median number of daily contacts stratified by demographic attributes. Estimates and their 95% confidence intervals were derived using bootstrap resampling (10,000 iterations).

Median daily contacts varied by participant characteristics in broadly consistent ways across countries. Bootstrap estimates in Figure 1 showed that, among adults aged 20 years and older, median contacts generally decreased with age. Participants with a university-level education tended to report more contacts than those with lower educational attainment. Contact counts also differed by employment status, with employed participants and students generally reporting more contacts than retired or unemployed participants. Median contacts increased with household size, and weekday reports were associated with higher median contacts than weekend reports.

**Fig 1.**
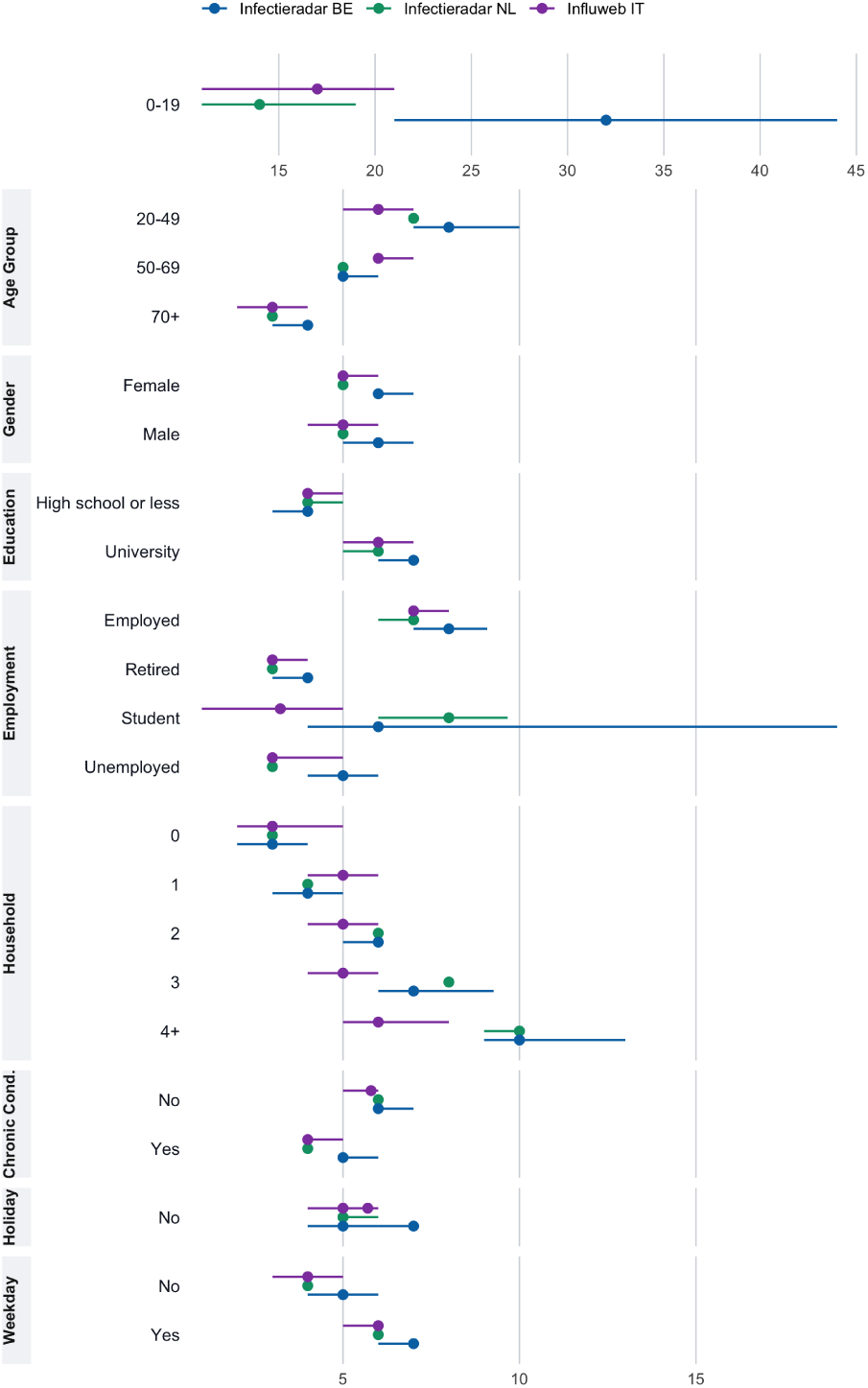
Median daily contacts by participant characteristics and status. Weighted median daily contacts by demographic characteristics for the Infectieradar platform in Belgium, Influweb in Italy, and the Infectieradar platform in the Netherlands, with 95% bootstrap-based confidence intervals. Note the 0–19 age group is shown with an independent x-axis scale.

Estimates for participants aged 0–19 were less stable than those for adult age groups, with substantially wider bootstrap confidence intervals and median contact distributions presented on an independent scale in Fig. 1. These unstable estimates were likely driven by small sample sizes in these strata, as shown in Table 2. For this reason, participants aged 0–19 years in all countries were excluded from subsequent analyses.

### 3.2 Exploring correlates of contact behavior

We fit negative binomial regression models to estimate correlates of the number of contacts of each adult participant, exploring social and demographic features, household size, day of the week, holiday period and reported symptoms. We applied post-stratification weights for age, sex, and day of week in model estimation, and we included participant-level random intercepts to account for repeated measures. Estimated coefficients are reported in Table 4. Further details on weighting and model specification are provided in Section 2.6.3. Weekday responses were associated with higher contact counts compared with weekends, consistent with increased work-related contacts during the week, in Italy (IRR = 1.374; 95% CI: 1.179–1.602) and the Netherlands (IRR = 1.049; 95% CI: 1.022–1.076), but not Belgium. Similarly, holiday periods were consistently associated with fewer contacts in all three countries, corresponding to reductions of approximately 20% in Belgium (IRR = 0.803; 95% CI: 0.705–0.915), 23% in Italy (IRR = 0.771; 95% CI: 0.694–0.856), and 9% in the Netherlands (IRR = 0.908; 95% CI: 0.889–0.926) compared to non-holiday periods.

**Table 4.** Negative binomial regression results (prioritizing temporal proximity of symptoms) for adult participants. Results from the negative binomial regression of contact counts from the Infectieradar platform in Belgium, Influweb in Italy, and the Infectieradar platform in the Netherlands, including random intercepts for participants. Estimates were obtained using post-stratification weights for age, sex, and day of week. The incidence rate ratio (IRR) is the exponentiated coefficient.

| Variable | BE |  |  | IT |  |  | NL |  |  |
| --- | --- | --- | --- | --- | --- | --- | --- | --- | --- |
|  | Coeff | IRR | P-value | Coeff | IRR | P-value | Coeff | IRR | P-value |
| Age group (ref: 50–69) |  |  |  |  |  |  |  |  |  |
| 20–49 | -0.128 | 0.880 | 0.279 | -0.230 | 0.795 | 0.047* | -0.050 | 0.951 | 0.015* |
| 70+ | 0.135 | 1.145 | 0.290 | -0.210 | 0.810 | 0.184 | -0.036 | 0.965 | 0.196 |
| Gender (ref: Female) | -0.160 | 0.853 | 0.074† | -0.047 | 0.955 | 0.658 | -0.116 | 0.891 | < 0.001*** |
| Education (ref: HS or less) | 0.442 | 1.556 | < 0.001*** | 0.088 | 1.092 | 0.369 | 0.023 | 1.024 | 0.171 |
| Employment (ref: Employed) |  |  |  |  |  |  |  |  |  |
| Student | – | – | – | -0.119 | 0.888 | 0.557 | – | – | – |
| Unemployed | -0.154 | 0.857 | 0.372 | -0.853 | 0.426 | < 0.001*** | -0.430 | 0.651 | < 0.001*** |
| Retired | -0.525 | 0.592 | < 0.001*** | -0.702 | 0.496 | < 0.001*** | -0.248 | 0.780 | < 0.001*** |
| Household Size | 0.143 | 1.153 | < 0.001*** | -0.016 | 0.984 | 0.474 | 0.145 | 1.156 | < 0.001*** |
| Chronic Condition (ref: False) | 0.115 | 1.122 | 0.252 | 0.095 | 1.099 | 0.358 | -0.056 | 0.945 | 0.001** |
| Holiday period (ref: False) | -0.219 | 0.803 | < 0.001*** | -0.261 | 0.771 | < 0.001*** | -0.097 | 0.908 | < 0.001*** |
| Survey day (ref: Weekend) | 0.056 | 1.057 | 0.507 | 0.318 | 1.374 | < 0.001*** | 0.048 | 1.049 | < 0.001*** |
| Symptoms (ref: None) |  |  |  |  |  |  |  |  |  |
| Symptomatic, no ILI | 0.146 | 1.157 | 0.231 | -0.032 | 0.968 | 0.734 | -0.009 | 0.991 | 0.540 |
| ILI | 0.064 | 1.066 | 0.774 | 0.288 | 1.334 | 0.101 | -0.058 | 0.944 | 0.020* |
| ILI with fever | -1.591 | 0.204 | < 0.001*** | 0.072 | 1.074 | 0.758 | -0.391 | 0.676 | < 0.001*** |

† $p < 0.10$ , \* $p < 0.05$ , \*\* $p < 0.01$ , \*\*\* $p < 0.001$

Employment status was also associated with contact counts. Retired participants reported fewer contacts than employed participants in all countries, with reductions of 41% in Belgium (IRR = 0.592; 95% CI: 0.454– 0.771), 50% in Italy (IRR = 0.496; 95% CI: 0.367–0.669), and 22% in the Netherlands (IRR = 0.780; 95% CI: 0.739–0.824). Unemployed participants reported 57% fewer contacts in Italy (IRR = 0.426; 95% CI: 0.302–0.602) and 35% fewer in the Netherlands (IRR = 0.651; 95% CI: 0.614–0.689).

In Italy (IRR = 0.795; 95% CI: 0.634–0.997) and the Netherlands (IRR = 0.951; 95% CI: 0.913–0.990), participants aged 20–49 reported fewer contacts than those aged 50–69. Gender differences were observed in the Netherlands, where males reported fewer contacts than females (IRR = 0.891; 95% CI: 0.861–0.921). In Belgium, the association was in the same direction but only marginally significant (IRR = 0.853; 95% CI: 0.716–1.015).

Household size was positively associated with contact counts in Belgium and the Netherlands. Each additional household member was associated with higher contact counts in Belgium (IRR = 1.153; 95% CI: 1.067–1.247) and the Netherlands (IRR = 1.156; 95% CI: 1.138–1.173).

Education level was associated with contact behavior only in Belgium. Adult participants with university education reported more contacts than those with high school education or less (IRR = 1.556; 95% CI: 1.262– 1.918). Chronic conditions were associated with lower contact counts only in the Netherlands, where those with a chronic condition reported fewer contacts than those without one (IRR = 0.945; 95% CI: 0.914–0.978). Regarding symptoms, when contact reports were linked by prioritizing temporal proximity of symptoms, symptom-related reductions in contact counts were observed in Belgium and the Netherlands. In Belgium, the most severe category, ILI with fever, was associated with fewer contacts compared with participants reporting no symptoms (IRR = 0.204; 95% CI: 0.083–0.500). In the Netherlands, ILI was associated with a moderate reduction of contacts (IRR = 0.944; 95% CI: 0.899–0.991), while ILI with fever was associated with 32% fewer contacts (IRR = 0.676; 95% CI: 0.619–0.731).

As an alternative specification, symptom status was defined by prioritizing symptom burden rather than temporal proximity, with detailed results presented in the Supporting Information. In Belgium, ILI with fever remained associated with fewer contacts (IRR = 0.547; 95% CI: 0.309–0.969). In the Netherlands, ILI was no longer significant while ILI with fever remained significantly associated with fewer contacts (IRR = 0.921; 95% CI: 0.874–0.971). Notably, symptomatic reports without ILI became associated with higher contact counts (IRR = 1.057; 95% CI: 1.032–1.082). Estimates for non-symptom covariates remained consistent.

Sensitivity analyses including minor participants produced similar model results. These analyses are presented and discussed in the Supporting Information.

### 3.3 Contact matrix comparison across studies

Contact matrices for adult participants in each study are shown in Figs. 2– 4, and matrix comparison metrics are reported in Table 5. Setting-specific matrices for home, work, and leisure contacts, together with the corresponding comparison metrics, are presented in the Supplementary Information.

**Fig 2.**
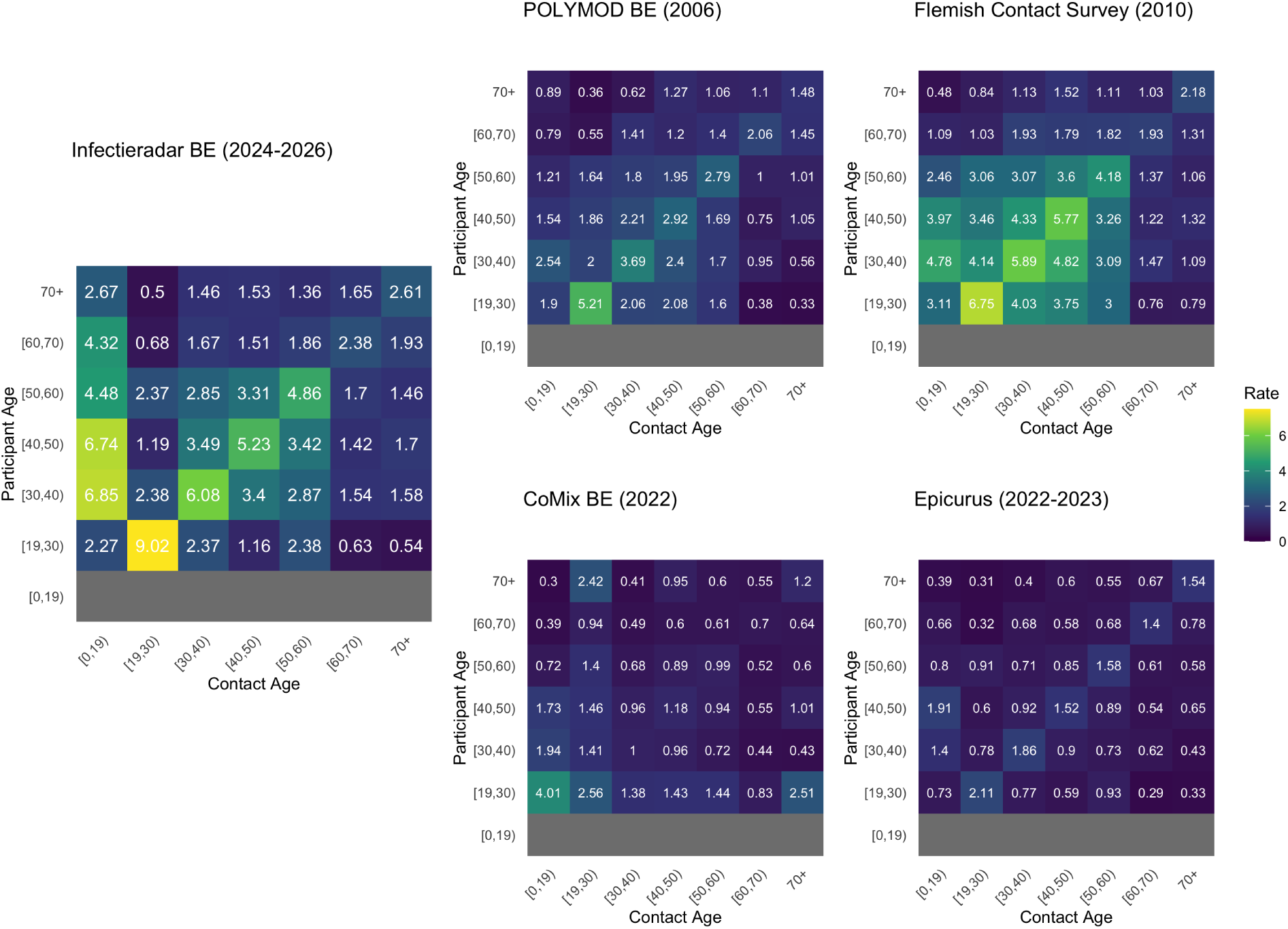
Belgian contact matrices for adult participants. Contact matrices comparing the Belgian Infectieradar matrix with Belgian comparator studies.

**Fig 3.**
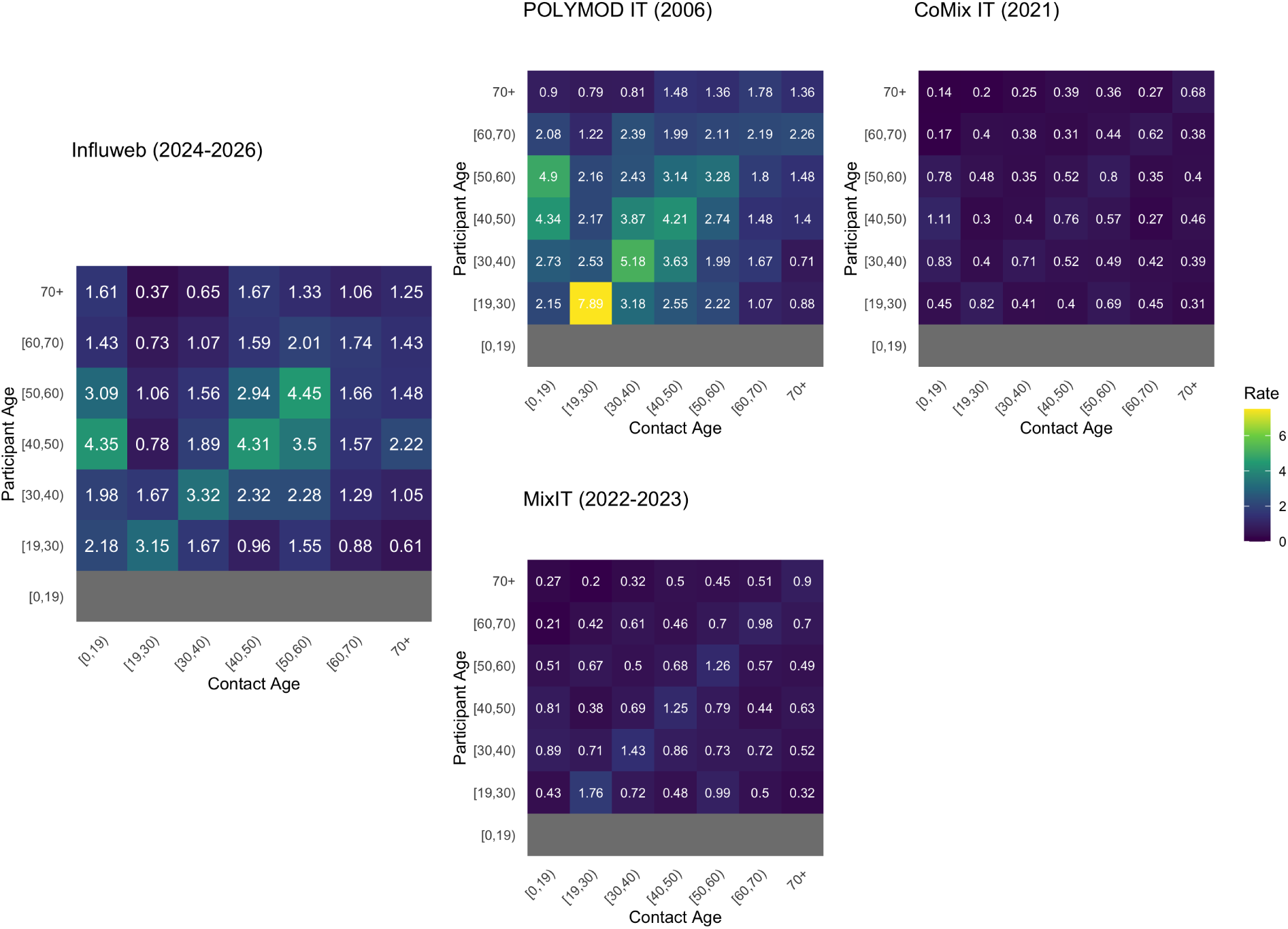
Italian contact matrices for adult participants. Contact matrices comparing the Influweb matrix with Italian comparator studies.

**Fig 4.**
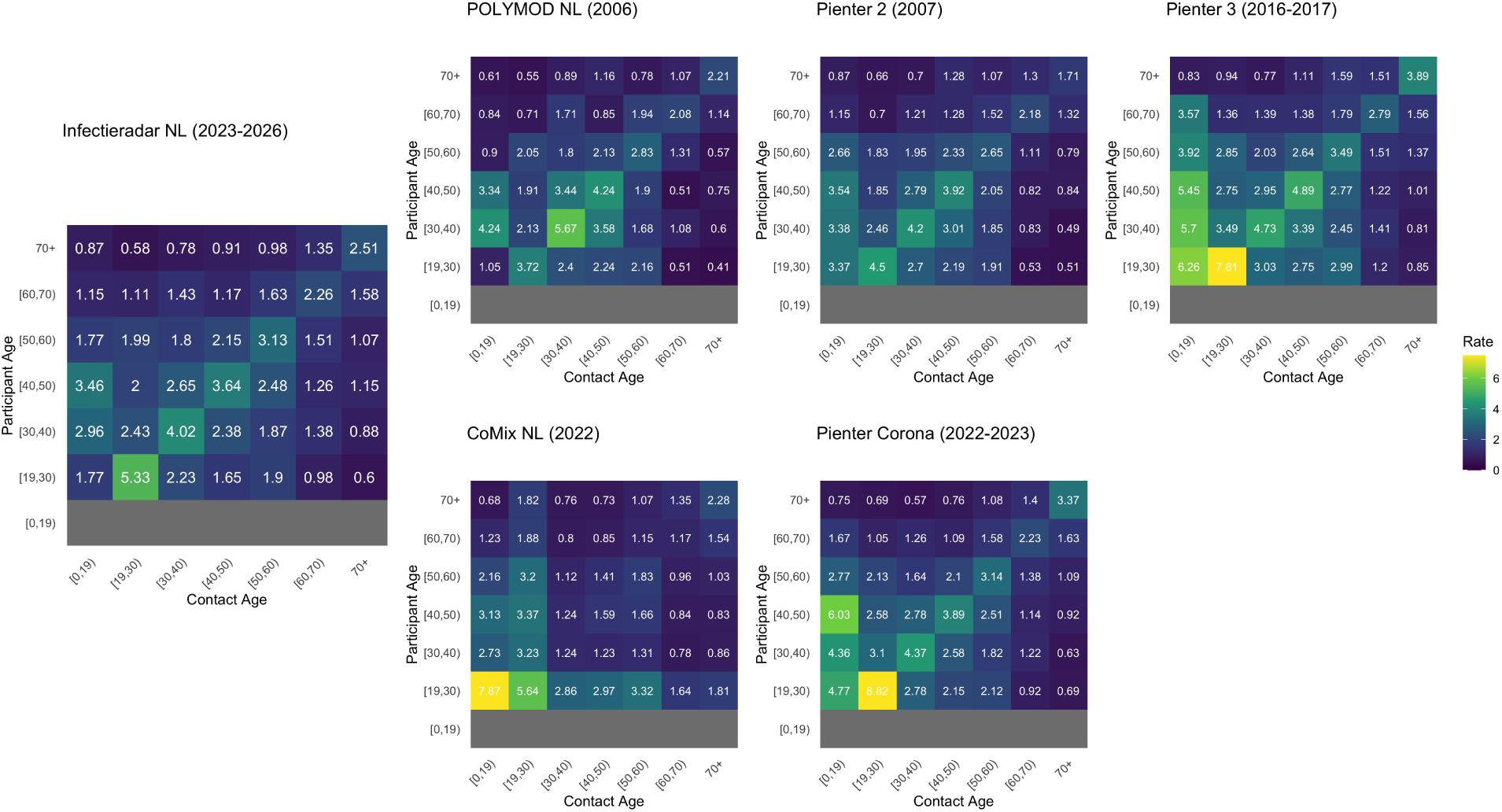
Dutch contact matrices for adult participants. Contact matrices comparing the Dutch Infectieradar matrix with Dutch comparator studies.

**Table 5.** Matrix comparison metrics by country. Cosine similarity and Sørensen–Dice Index range from 0 to 1, with larger values indicating greater similarity to the country-specific reference matrix. Frobenius norm measures absolute matrix distance, with smaller values indicating greater similarity

| Comparison metric | Cosine similarity | Sørensen–Dice index | Frobenius norm |
| --- | --- | --- | --- |
| **Belgium** (reference: Infectieradar BE) |  |  |  |
| POLYMOD BE | 0.934 | 0.721 | 10.995 |
| FCS | 0.929 | 0.839 | 7.870 |
| CoMix 2022 BE | 0.756 | 0.489 | 15.841 |
| Epicurus | 0.957 | 0.472 | 15.456 |
| **Italy** (reference: Influeweb) |  |  |  |
| POLYMOD IT | 0.928 | 0.821 | 7.300 |
| CoMix 2021 IT | 0.949 | 0.407 | 10.558 |
| MixIT | 0.930 | 0.530 | 9.380 |
| **Netherlands** (reference: Infectieradar NL) |  |  |  |
| POLYMOD NL | 0.964 | 0.882 | 3.772 |
| PIENTER 2 | 0.979 | 0.916 | 2.841 |
| PIENTER 3 | 0.962 | 0.830 | 7.794 |
| CoMix 2022 NL | 0.832 | 0.780 | 8.472 |
| PIENTER Corona | 0.966 | 0.890 | 5.806 |

In Belgium, cosine similarity was high for Epicurus, POLYMOD, and the FCS, indicating broadly similar age-mixing structure to the Infectieradar BE matrix. However, the Sørensen–Dice index and Frobenius norm suggest that the FCS showed the strongest overlap in contact volume and smaller absolute differences in contact intensity. CoMix showed weaker agreement across metrics, indicating greater differences from the Infectieradar BE matrix in both structure and contact distribution.

In Italy, cosine similarity was high across all comparisons, indicating that cells with relatively high or low contact rates were generally consistent between the Influweb matrix and each comparator matrix. The Sørensen–Dice index was higher for POLYMOD and MixIT than for CoMix, suggesting greater overlap in the distribution of contact volume, while the higher Frobenius norm for CoMix indicated larger absolute differences in contact intensity.

In the Netherlands, the comparator matrices showed high agreement with the Infectieradar NL matrix across all three metrics, indicating similar age-mixing structure, substantial overlap in contact volume, and relatively small absolute differences in contact intensity. CoMix showed the lowest relative agreement among the Dutch comparators, although similarity remained moderate to high.

In the correlation plots, both the BE (Fig. 5) and NL (Fig. 7) Infectieradar platforms showed strong agreement with most comparator studies, with the exception of CoMix, which showed weak agreement in both countries. Influweb in Italy (Fig. 6) showed more moderate but consistent correlations across comparator datasets, indicating broadly similar relative mixing patterns but greater variation in individual age-pair values than observed for the Infectieradar comparisons.

**Fig 5.**
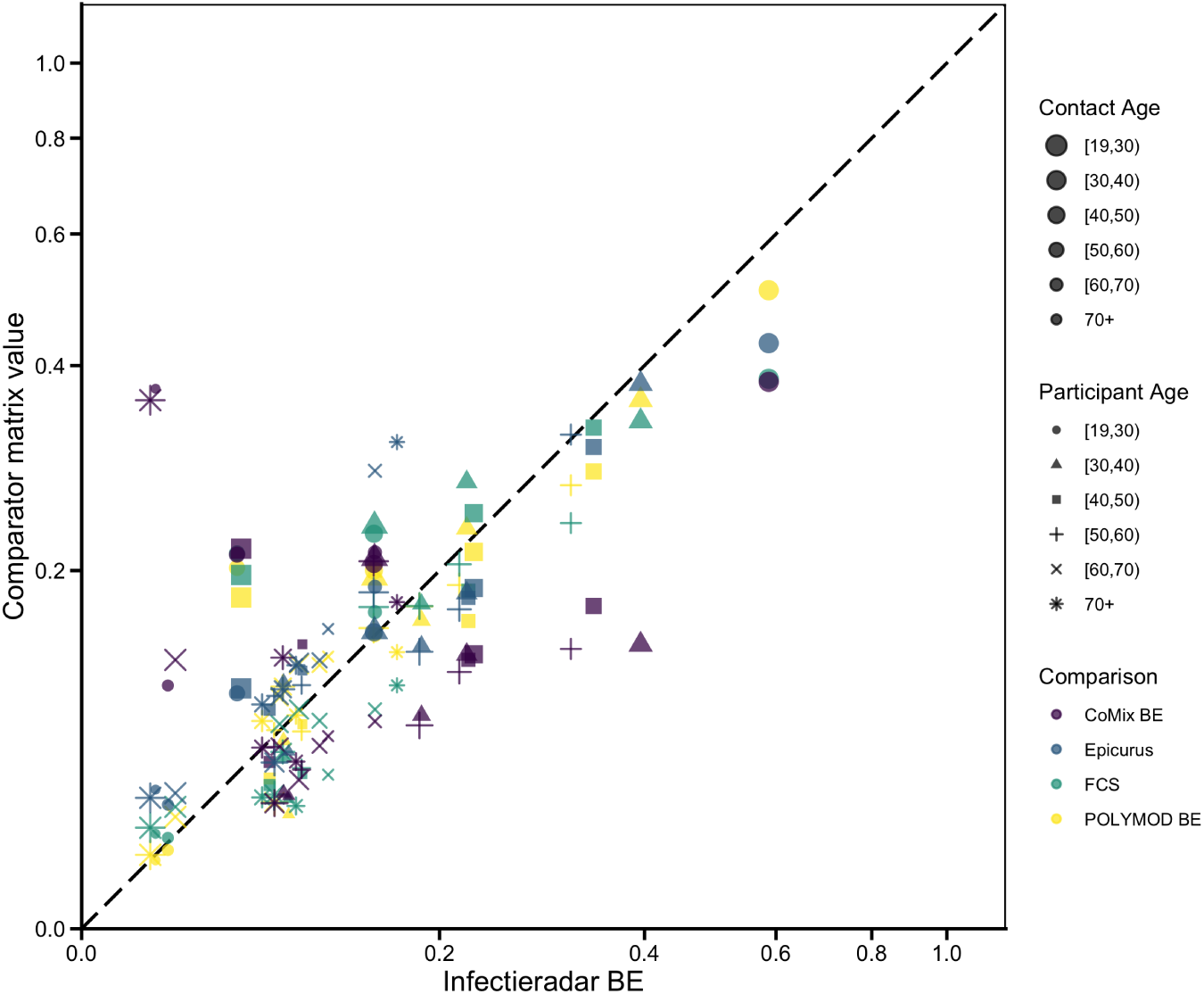
Belgian square contact matrix correlations. The plot shows normalized age-pair contact values from the Belgian Infectieradar square contact matrix compared with Belgian comparator studies. Color indicates the comparator study, point shape indicates participant age group, and point size indicates contact age group. Values are scaled so that the dominant eigenvalue equals one. Points closer to the diagonal indicate stronger agreement between the two matrices.

**Fig 6.**
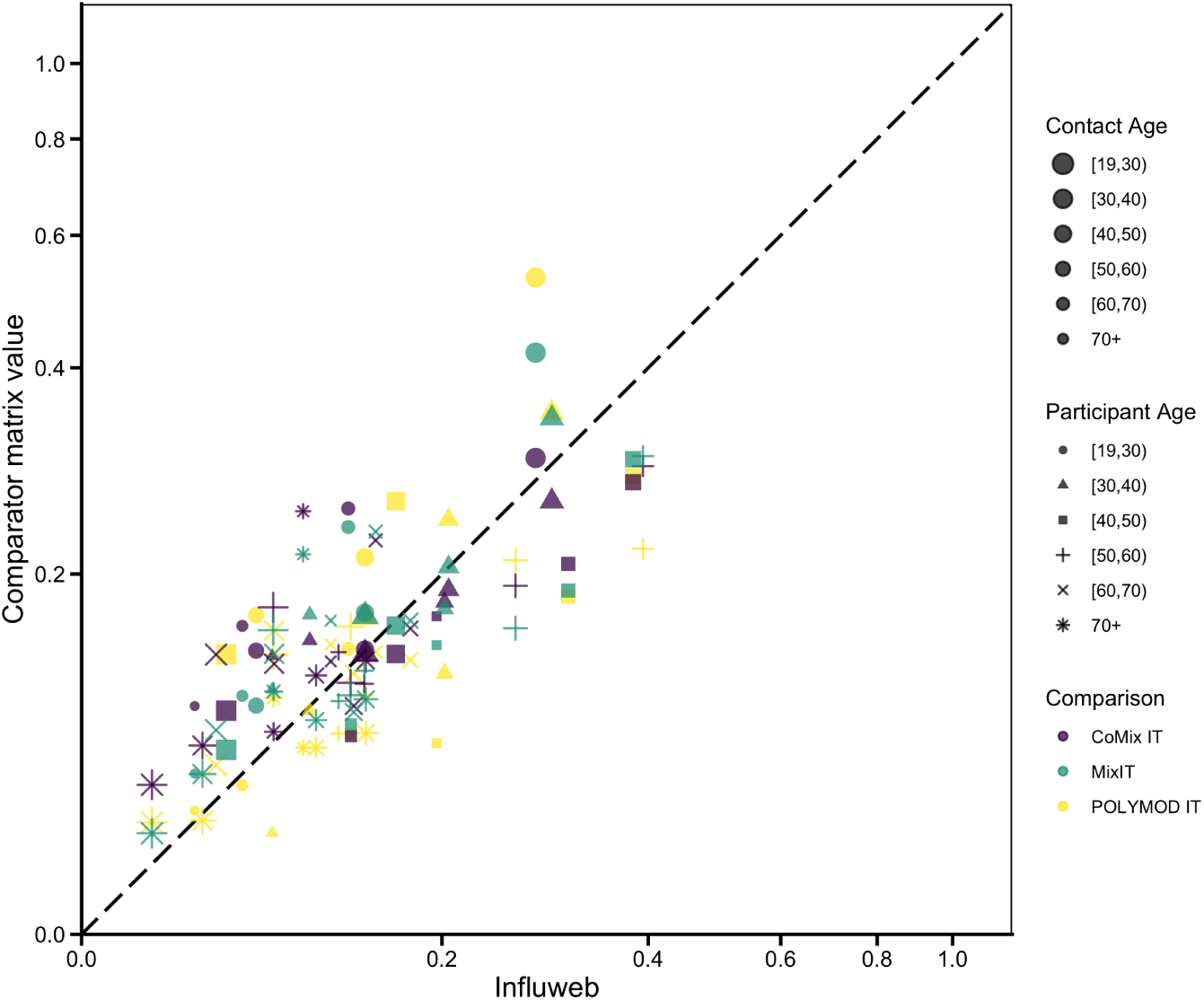
Italian square contact matrix correlations. The plot shows normalized age-pair contact values from the Influweb square contact matrix compared with Italian comparator studies. Color indicates the comparator study, point shape indicates participant age group, and point size indicates contact age group. Values are scaled so that the dominant eigenvalue equals one. Points closer to the diagonal indicate stronger agreement between the two matrices.

**Fig 7.**
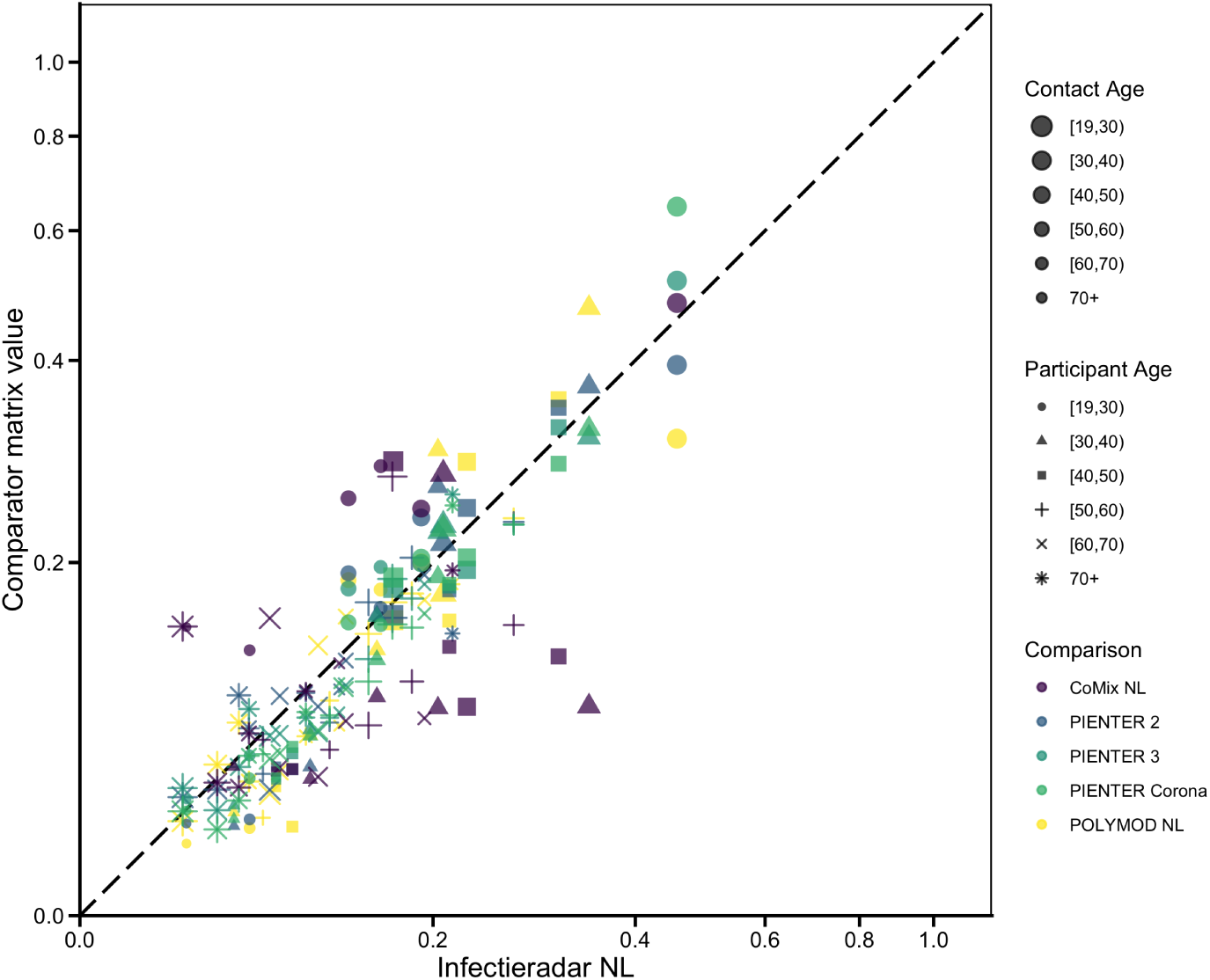
Dutch square contact matrix correlations. The plot shows normalized age-pair contact values from the Dutch Infectieradar square contact matrix compared with Dutch comparator studies. Color indicates the comparator study, point shape indicates participant age group, and point size indicates contact age group. Values are scaled so that the dominant eigenvalue equals one. Points closer to the diagonal indicate stronger agreement between the two matrices.

**Fig 8.**
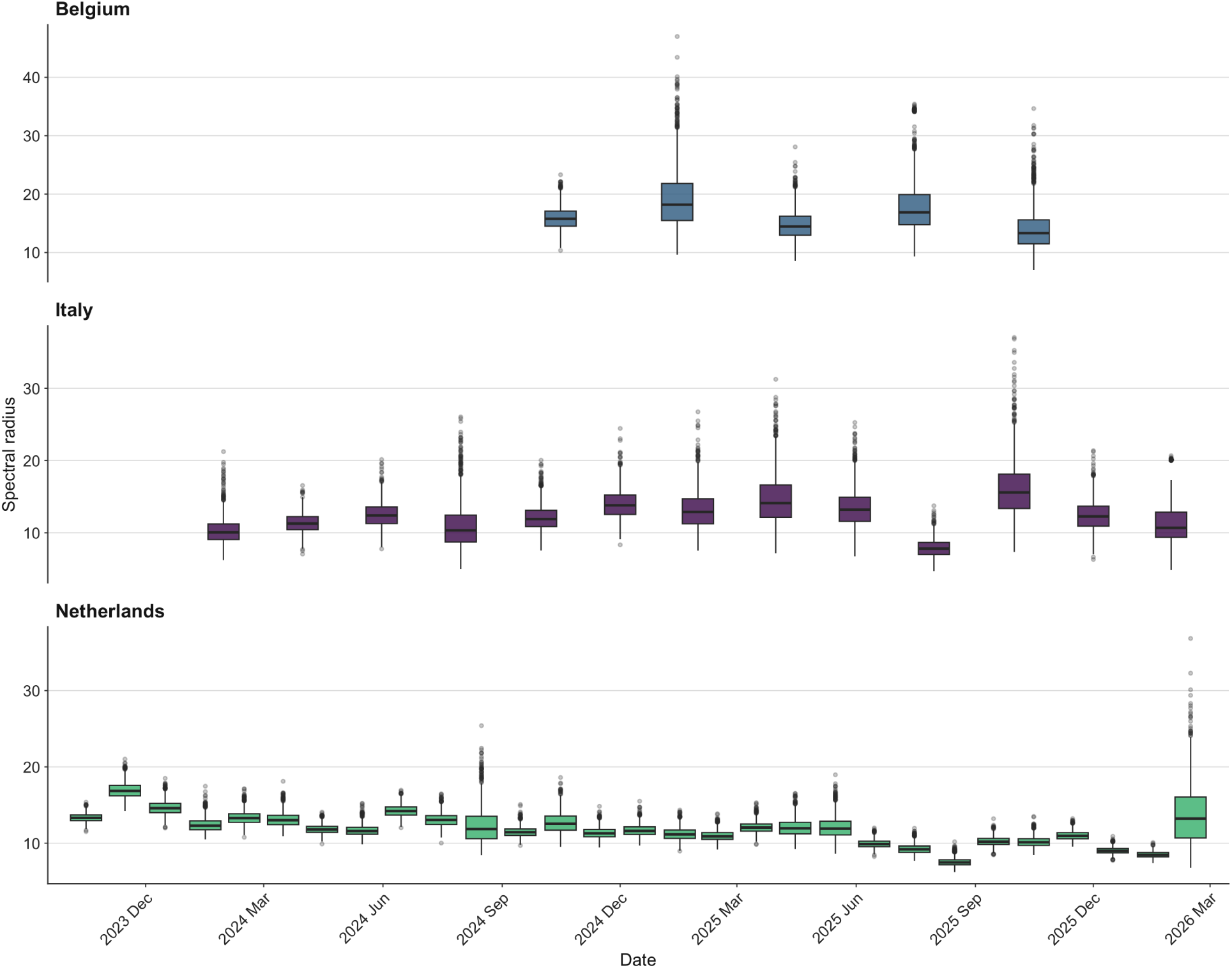
Distribution of the spectral radius of bootstrapped adult contact matrices over time by country. Each boxplot shows the distribution of dominant eigenvalues across bootstrapped symmetric adult contact matrices for a given date grouping. Date groupings differ by country: Belgium is grouped by season-year, Italy by two-month periods, and the Netherlands by monthly periods.

For Italy and the Netherlands, we also repeated the comparisons across studies for contact matrices including minor participants, with results reported in the Supporting Information. The comparison metrics remained broadly stable relative to the adult-only analyses, indicating that the overall conclusions were not sensitive to excluding minors. In most comparisons, similarity increased after the inclusion, due to the strong minor-to-minor contact patterns visible in the full-age matrices.

Taken together, these comparisons suggest that the participatory surveillance platforms capture the main age-mixing structure observed in established contact surveys. Differences in absolute intensity, reflected in the Frobenius distances, are expected given differences in survey mode, contact definition, sampling strategy, and social context, and should not be interpreted as invalidating the participatory data source.

### 3.4 Distributions of contact intensity over time

The distribution of contact intensity, here represented by the spectral radii, varied over time within each country, demonstrating that repeated participatory contact surveys can capture temporal changes in transmission-relevant social mixing. The magnitude and variability of these estimates differed by country, partly reflecting differences in sample size, date grouping, and study duration. The Netherlands, with the largest number of observations and monthly grouping, provided the highest temporal resolution, whereas Belgium and Italy required broader time groupings to maintain stable adult contact matrices. These results illustrate the main added value of the participatory surveillance approach: once a platform is established, it can generate repeated, internally consistent summaries of contact patterns over time.

## 4 Discussion

In this study, we explored whether participatory surveillance platforms can collect valid, meaningful social contact data relevant to infectious disease transmission. To this end, contact matrices derived from three InfluenzaNet platforms were compared with large, representative contact surveys and found to be broadly consistent. The Dutch Infectieradar matrix showed the closest overall agreement with comparator datasets, while the Italian Influweb and Belgian Infectieradar matrices showed high structural similarity but less agreement in absolute contact intensity and volume overlap.

We consider agreement in relative age-mixing structure more informative than exact agreement in absolute contact rates, which are sensitive to survey design, sample composition, and the social context of data collection. For example, the Italian CoMix data was collected while varying social-distancing measures were in place ^33^. Relative age-mixing structure, by contrast, is central to age-structured epidemic models and was the dimension on which the InfluenzaNet and comparator matrices agreed most strongly. Matrix-similarity metrics also remained stable or increased when minor participants were included, shown in the Supporting Information, while expected model associations with weekends, holidays, and employment were consistent with previous contact studies ^8;28;46;47;65;74^. Together, these findings support the validity of the contact patterns captured by the platforms. Having established this validity, the main advantage of participatory surveillance lies in its ability to update contact matrices repeatedly and provide current data for outbreak modeling.

The longitudinal spectral-radius analysis illustrates this distinctive advantage. InfluenzaNet and similar platforms draw on their established infrastructure and engaged participant communities to integrate new survey types and conduct repeated rounds of data collection at relatively low marginal cost. Under fixed epidemiological assumptions, temporal variation in the spectral radius summarizes changes in the transmission-relevant component of social mixing within each platform. The scenario-based *R*_0_ distributions presented in the Supporting Information similarly shows how transmission potential associated with contact patterns vary across periods and epidemiological contexts. Though these analyses should not be interpreted as direct estimates of infectiousness, they provide summaries of transmission-relevant contact patterns under a common epidemiological framework. These variations within and across studies underscore the limitations of treating contact matrices as static inputs. By showing that contact measurement can be repeated within existing surveillance infrastructure, this study demonstrates a practical way to maintain a living source of regularly updated contact information for epidemic models.

A further major contribution of participatory surveillance is the ability to link contact behavior with information collected from the same participants across multiple surveys covering a broad range of topics. InfluenzaNet platforms routinely collect sociodemographic characteristics, vaccination status, comorbidities, symptom histories, risk perception, and health-related behaviors, while complementary survey modules can be launched in response to emerging public health topics. This constitutes a unique strength of participatory platforms, which produce longitudinal observations across a wide range of participant-level characteristics that are rarely available through other data-collection modalities. This data can be assembled without creating ad hoc cohorts or initiating separate recruitment for each survey type.

Individual-level linkage between symptom histories and contact behavior, in particular, remains rare and difficult to achieve in social contact research. Our results revealed a nuanced association between these measures. When contact reports were linked to the most recent prior symptom survey, reductions in contact counts were observed among participants with more severe symptom profiles. In Belgium, ILI with fever was associated with fewer reported contacts, while in the Netherlands both ILI and ILI with fever were associated with contact reductions. In the alternative symptom specification, which prioritized the symptom report in the prior 30 days with the highest symptom burden rather than temporal proximity, ILI with fever remained associated with fewer contacts in Belgium and the Netherlands, supporting an interpretation that those with a more severe symptom status reduced their social mixing. However, ILI was no longer associated with fewer contacts in the Netherlands, while symptomatic reports that did not fulfill the ILI definition were associated with higher contact counts. This suggests that individuals reporting mild symptoms may remain socially active, or higher-contact individuals may report more symptoms. Although we cannot comment on the direction of this association from our observational study, this result is notable.

Few studies have directly linked social contact behavior with symptom status, though our findings are consistent with the available evidence indicating that the association between symptoms and social behavior depends on study design, symptom severity, and timing of measurement. An analysis of aggregated population trends in weekly contacts and respiratory symptoms reported that peaks in contacts coincided with peaks in symptoms ^75^, supporting the possibility that periods or groups with higher contact activity may also report greater symptom burden. In contrast, reduced contacts have been observed among symptomatic individuals recruited from antiviral distribution centers ^76^, a study sample more likely to present with severe illness, and in the Epistorm-Mix study, where feeling sick on the contact-diary day was associated with fewer reported contacts ^77^. Together with our findings, this evidence suggests that mild or population-level symptom burden may coexist with sustained social activity, while more severe illness is more consistently associated with contact reduction.

More broadly, the capacity to link repeated contact reports with participant characteristics positions InfluenzaNet platforms to investigate disparities in exposure and disease burden, an increasingly important direction of infectious disease research ^15–17^. This linked longitudinal data could reveal whether exposure inequalities widen or narrow over time, differ across population groups or countries, or change during public health interventions. Such analyses would extend the study of contact patterns beyond age alone and help identify groups whose social circumstances affect their exposure. Linkage with biological sampling, as piloted through participant self-swabbing in the Netherlands ^78^, could also connect behavioral, clinical, and pathogen information. This flexibility allows participatory surveillance systems to respond to changing public health priorities while maintaining a consistent longitudinal core.

While these findings highlight the value of participatory surveillance, our approach is not without limitations. InfluenzaNet’s participants are self-selected, which may overrepresent health-conscious individuals ^38^. Post-stratification weighting can reduce measured demographic imbalances but cannot eliminate selection bias. As such, the results should be interpreted as patterns observed within an engaged surveillance cohort rather than as nationally representative estimates. The observational design precludes causal inference, and self-reported contacts and symptoms may be affected by recall or social desirability bias, though these effects were likely limited by the short recall windows and relatively stable public health context during data collection. Survey fatigue is a concern in longitudinal designs, but response rates stabilized after survey launch. Recruitment of children, through adults reporting on behalf of minors, remains a particular challenge thus the number of younger participants was limited, reducing our ability to characterize contact patterns involving younger age groups. Nevertheless, sensitivity analyses including and excluding minor participants produced broadly consistent results in the matrix comparisons and regression analyses, suggesting that the main findings were robust to this limitation.

Despite these limitations, InfluenzaNet platforms have a well-established record of producing reliable epidemiological signals. More than a decade of evidence shows that their citizen-reported ILI estimates track established sentinel surveillance systems and can provide a complementary view of population health ^37;38;40;45^. Participatory systems have also provided timely information during periods when traditional surveillance systems were delayed or under pressure ^79–81^. Our findings extend this track record to social contact data and support the development of a participatory European social contact observatory.

InfluenzaNet provides a concrete foundation for this next step. The network already operates in 12 countries and draws on infrastructure and collaborations supported by EU-funded initiatives, providing an existing institutional and funding base from which the observatory could be developed. Cross-country collaboration allows common survey instruments to be deployed across national platforms, producing harmonized data that is more directly comparable across borders^37;45^. One central function of such an observatory would be to establish and maintain context-specific reference baselines for social mixing. Rather than relying on a fixed representation of population behavior, repeated collection could characterize expected seasonal, demographic, and country-specific variation in contact volume and structure. Departures from these reference patterns could then be interpreted in relation to outbreaks, interventions, changes in risk perception, or other major social disruptions. With adequate temporal resolution, contact indicators could become a behavioral signal that complements syndromic surveillance by identifying changes in transmission-relevant behavior before they are fully reflected in case counts, health-care use, or other epidemiological outcomes.

An operational European social contact observatory would translate this infrastructure into a sustained public health resource. A regularly updated dashboard and data feed could provide summaries of contact behavior that public health institutes and epidemic-modeling groups can use to interpret epidemiological trends alongside changes in social mixing, update transmission scenarios, and help assess behavioral responses to emerging health threats and public health interventions. By making social contact behavior a routine component of surveillance, public health institutions would enter future crises with established baselines and the capacity to monitor behavioral change as it unfolds, ultimately improving European pandemic preparedness.

## 5 Data availability

The individual-level data used in this study from the InfluenzaNet platforms cannot be shared publicly because they contain information that could compromise participant privacy. Requests for access to the individual-level data may be directed to the authors.

Aggregated data products derived from these data, analysis code, and figure-generation scripts, are available on GitHub and archived on Zenodo (DOI 10.5281/zenodo.22074901).

The comparator datasets are catalogued at https://socialcontactdata.org/data/ and deposited on Zenodo, with the exception of the Pienter2 data, which is not publicly available at the time of publication.

## 7 Author contribution

All authors contributed to the design of the study. PC, LH, DP, AJVH participated in the design of the original survey. Data was processed and prepared for analyses by NG, LH, KNK, and MM. KNK performed the statistical analysis, with oversight from PC, DP, MM and NG. The first draft of the manuscript was produced by KNK and all authors reviewed, edited, and approved the final version.

## Supporting information

Supplementary analyses and interpretation supporting the results presented in the main text.

## Data Availability

The individual-level data used in this study from the InfluenzaNet platforms cannot be shared publicly because they contain information that could compromise participant privacy. Requests for access to the individual-level data may be directed to the authors.
Aggregated data products derived from these data, analysis code, and figure-generation scripts, are available on GitHub and archived on Zenodo (DOI 10.5281/zenodo.22074901).
The comparator datasets are catalogued at https://socialcontactdata.org/data/ and deposited on Zenodo, with the exception of the Pienter2 data, which is not publicly available at the time of publication.

https://doi.org/10.5281/zenodo.22074902

## 8 Supporting information

**S1 Table. Negative binomial regression results prioritizing symptom burden, for adult participants.** Results from the negative binomial regression of contact counts from the Infectieradar platform in Belgium, Influweb in Italy, and the Infectieradar platform in the Netherlands, including random intercepts for participants. Estimates were obtained using post-stratification weights for age, sex, and day of week. The incidence rate ratio (IRR) is the exponentiated coefficient.

**S2 Table. All-age contact matrix comparison metrics for Italy and the Netherlands.** Italy comparisons use Influweb as the reference matrix. Netherlands comparisons use Infectieradar as the reference matrix. Higher cosine similarity and Sørensen–Dice index indicate greater similarity; lower Frobenius norm indicates smaller absolute matrix differences.

**S1 Fig. All-age Italy contact matrices across studies.** Mean bootstrap contact matrices are shown for Influweb, POLYMOD IT, CoMix IT, and MixIT. All panels use the same color scale.

**S2 Fig. All-age Netherlands contact matrices across studies.** Mean bootstrap contact matrices are shown for Infectieradar NL, POLYMOD NL, PIENTER 2, PIENTER 3, CoMix NL, and PIENTER Corona. All panels use the same color scale.

**S3 Fig. All-age Italy matrix correlation plots.** Each point represents one matrix cell after dominant-eigenvalue normalization. Influweb is the reference matrix and the dashed line is the identity line.

**S4 Fig. All-age Netherlands matrix correlation plots.** Each point represents one matrix cell after dominant-eigenvalue normalization. Infectieradar NL is the reference matrix and the dashed line is the identity line.

**S3 Table. Negative binomial regression results prioritizing temporal proximity of symptoms, all ages.** Results from the negative binomial regression of contact counts from the Infectieradar platform in Belgium, Influweb in Italy, and the Infectieradar platform in the Netherlands, including random intercepts for participants. These models include all ages in the sample. Estimates were obtained using post-stratification weights for age, sex, and day of week. The incidence rate ratio (IRR) is the exponentiated coefficient. The IT model includes a zero-inflation intercept.

**S4 Table. Negative binomial regression results prioritizing symptom burden, all ages.** Results from the negative binomial regression of contact counts from the Infectieradar platform in Belgium, Influweb in Italy, and the Infectieradar platform in the Netherlands, including random intercepts for participants. These models include all ages in the sample. Estimates were obtained using post-stratification weights for age, sex, and day of week. The incidence rate ratio (IRR) is the exponentiated coefficient. The IT model includes a zero-inflation intercept.

**S5 Fig. Italy setting-specific contact matrices for adults across studies.** Mean bootstrapped home, work, and leisure contact matrices are shown for Italian studies included in the setting-specific comparison. **S6 Fig. Netherlands setting-specific contact matrices for adults across studies.** Mean boot-strapped home, work, and leisure contact matrices are shown for Dutch studies included in the setting-specific comparison.

**S7 Fig. Belgium setting-specific contact matrices across studies.** Mean bootstrapped home, work, and leisure contact matrices are shown for Belgian studies included in the setting-specific comparison.

**S5 Table. Setting-specific contact matrix comparison metrics by country.** Metrics compare each setting-specific matrix with the country-specific participatory surveillance reference matrix.

**S8 Fig. Belgian bootstrapped** *R*_0_ **distributions across studies.** Distributions of derived *R*_0_ values based on Belgian contact matrices from Infectieradar BE and comparator studies.

**S9 Fig. Italian bootstrapped** *R*_0_ **distributions across studies.** Distributions of derived *R*_0_ values based on Italian contact matrices from Influweb and comparator studies.

**S10 Fig. Dutch bootstrapped** *R*_0_ **distributions across studies.** Distributions of derived *R*_0_ values based on Dutch contact matrices from Infectieradar NL and comparator studies.

