## Supplementary analyses and interpretation supporting the results presented in the main text. for "Towards a participatory European social contact observatory"

This supporting information provides additional methodological details, sensitivity analyses, and supplementary results for the main manuscript. Sections are organized to follow the order in which related analyses are introduced in the main text.

#### 1 Post-stratification weights

Post-stratification weights were applied to align the study samples with the corresponding reference populations, defined as the Belgian, Italian, or Dutch general population in the year of data collection. Weights accounted for differences in the age and sex distribution of the survey sample relative to the reference population, as well as differences in the distribution of responses between weekdays and weekends.

First, we accounted for differences between the age and sex distribution of the survey sample and the reference population, a common practice in survey analysis[1].

Let $k = (s, a)$ denote a strata defined by sex $s \in \{M, F\}$ and age $a \in \{0{-}9, 10{-}19, \ldots, 90{-}99\}$. Weights were calculated by taking the ratio of the proportion in stratum $k$ of the reference country's population in a given survey year to the proportion of the study sample in the same stratum:

$$w_k = \frac{P_k/P}{N_k/N},$$

where $P$ represents the country's population and $N$ the sample. Population counts were obtained from national demographic data[2–4].

Then we adjusted for differences in survey response timing. Contact patterns differ by day of the week, with higher rates often seen on weekdays at work and school. To ensure that weekday and weekend responses reflect their share of the calendar week, we apply day-of-week weights:

$$w_{day.of.week} = \frac{5/7}{N_{weekday}/N} \quad \text{or} \quad \frac{2/7}{N_{weekend}/N},$$

where $N_{weekend(weekday)}$ is the number of surveys returned on the weekend(weekday), and $N$ is the total number of surveys.

The final weight for participant $p$ is the product of the age-sex and day-of-week weights: $w_p = w_k \times w_{day.of.week}$. To limit the influence of observations with very large post-stratification adjustments, final weights were capped at three before analysis[5]. This affected only a small number of observations, and sensitivity analyses showed consistent results with and without capped weights.

The weights were applied to the Influweb and both Infectieradar samples in all analyses, including contact matrix construction, regression modeling, and estimation of median daily contacts by participant characteristics. The same weighting approach was also applied when constructing contact matrices from comparator datasets (e.g., POLYMOD, CoMix) to ensure consistency and comparability across studies.

### 2 Sensitivity analyses

#### 2.1 Alternative symptom matching definition

Each contact survey entry was linked to symptom reports from the same participant. Because participants could submit multiple weekly symptom surveys within the 30-day window before a contact survey, symptom status was assigned using two matching approaches. The main-text model used the temporal proximity approach, selecting the symptom survey submitted closest in time before the contact survey, which often occurred on the same day. As a sensitivity analysis, Table S1 reports the symptom burden approach, which selected the symptom survey with the highest number of reported symptoms within the same 30-day window.

Compared with the temporal proximity model in the main text, the symptom burden model yielded broadly consistent results. In Belgium, ILI with fever remained associated with fewer contacts (IRR = 0.547; 95% CI: 0.309–0.969). In the Netherlands, ILI was no longer significantly associated with contact counts, while ILI with fever remained associated with fewer contacts (IRR = 0.921; 95% CI: 0.874–0.971). Symptomatic reports not meeting the ILI definition were associated with higher contact counts in the Netherlands (IRR = 1.057; 95% CI: 1.032–1.082). No symptom category was significantly associated with contact counts in Italy. Estimates for non-symptom covariates remained similar to those from the temporal proximity model.

#### 2.2 Contact matrix comparison across studies using all ages

As a sensitivity analysis of the exclusion of minor participants in the main-text analyses, we constructed the all-age contact matrices for the Italian and Dutch social contact datasets, following the procedure outlined in the main text (bootstrap sampling, post-stratification weighting, and reciprocity correction). We compared all-age contact matrices from the Italian and Dutch participatory surveillance datasets with all available

| Variable | BE Coeff | BE IRR | BE P-value | IT Coeff | IT IRR | IT P-value | NL Coeff | NL IRR | NL P-value |
|---|---|---|---|---|---|---|---|---|---|
| Age group (ref: 50–69) | | | | | | | | | |
| 20–49 | -0.136 | 0.873 | 0.256 | -0.229 | 0.795 | 0.048* | -0.058 | 0.943 | 0.005** |
| 70+ | 0.139 | 1.149 | 0.279 | -0.208 | 0.813 | 0.191 | -0.032 | 0.969 | 0.247 |
| Gender (ref: Female) | -0.156 | 0.856 | 0.082† | -0.050 | 0.951 | 0.636 | -0.111 | 0.895 | $< 0.001^{***}$ |
| Education (ref: HS or less) | 0.446 | 1.562 | $< 0.001^{***}$ | 0.083 | 1.086 | 0.401 | 0.022 | 1.023 | 0.193 |
| Employment (ref: Employed) | | | | | | | | | |
| Retired | -0.520 | 0.594 | $< 0.001^{***}$ | -0.708 | 0.492 | $< 0.001^{***}$ | -0.245 | 0.782 | $< 0.001^{***}$ |
| Student | – | – | – | -0.120 | 0.887 | 0.556 | – | – | – |
| Unemployed | -0.124 | 0.884 | 0.479 | -0.860 | 0.423 | $< 0.001^{***}$ | -0.432 | 0.650 | $< 0.001^{***}$ |
| Household Size | 0.148 | 1.159 | $< 0.001^{***}$ | -0.015 | 0.985 | 0.516 | 0.144 | 1.155 | $< 0.001^{***}$ |
| Chronic Condition (ref: False) | 0.126 | 1.134 | 0.215 | 0.097 | 1.101 | 0.349 | -0.060 | 0.942 | 0.001** |
| Holiday period (ref: False) | -0.222 | 0.801 | $< 0.001^{***}$ | -0.257 | 0.773 | $< 0.001^{***}$ | -0.098 | 0.907 | $< 0.001^{***}$ |
| Survey day (ref: Weekend) | 0.065 | 1.067 | 0.439 | 0.317 | 1.373 | $< 0.001^{***}$ | 0.047 | 1.048 | $< 0.001^{***}$ |
| Symptoms (ref: None) | | | | | | | | | |
| Symptomatic, no ILI | 0.066 | 1.069 | 0.499 | 0.020 | 1.020 | 0.772 | 0.056 | 1.057 | $< 0.001^{***}$ |
| ILI | 0.189 | 1.208 | 0.245 | -0.008 | 0.992 | 0.948 | 0.016 | 1.017 | 0.366 |
| ILI with fever | -0.604 | 0.547 | 0.039* | 0.111 | 1.118 | 0.421 | -0.082 | 0.921 | 0.002** |

† $p < 0.10$, * $p < 0.05$, ** $p < 0.01$, *** $p < 0.001$

**Table S1. Negative binomial regression results prioritizing symptom burden, for adult participants.** Results from the negative binomial regression of contact counts from the Infectieradar platform in Belgium, Influweb in Italy, and the Infectieradar platform in the Netherlands, including random intercepts for participants. Estimates were obtained using post-stratification weights for age, sex, and day of week. The incidence rate ratio (IRR) is the exponentiated coefficient.

country-specific comparator studies. Influweb was used as the reference for Italy, and Infectieradar was used as the reference for the Netherlands.

Belgian Infectieradar was not included in this sensitivity analysis because too few observations were available among minor participants.

The matrices are presented in Figs. S1 and S2. The quantitative matrix comparison metrics, described in the main text, are shown in Table S2, and the correlation plots are shown in Figs. S3 and S4. The comparison metrics remained broadly stable relative to the adult-only analyses. In most comparisons, similarity increased after including minors, consistent with the strong minor-to-minor contact patterns visible in the all-age matrices.

| Country | Comparator | Cosine similarity | Sørensen–Dice index | Frobenius norm |
|---|---|---|---|---|
| Italy | POLYMOD IT | 0.949 | 0.838 | 7.295 |
| | CoMix IT | 0.838 | 0.390 | 17.398 |
| | MixIT | 0.951 | 0.526 | 14.637 |
| Netherlands | POLYMOD NL | 0.971 | 0.873 | 4.691 |
| | PIENTER 2 | 0.981 | 0.886 | 4.529 |
| | PIENTER 3 | 0.988 | 0.879 | 5.706 |
| | CoMix NL | 0.933 | 0.793 | 7.093 |
| | PIENTER Corona | 0.929 | 0.854 | 18.221 |

**Table S2. All-age contact matrix comparison metrics for Italy and the Netherlands.** Italy comparisons use Influweb as the reference matrix. Netherlands comparisons use Infectieradar as the reference matrix. Higher cosine similarity and Sørensen–Dice index indicate greater similarity; lower Frobenius norm indicates smaller absolute matrix differences.

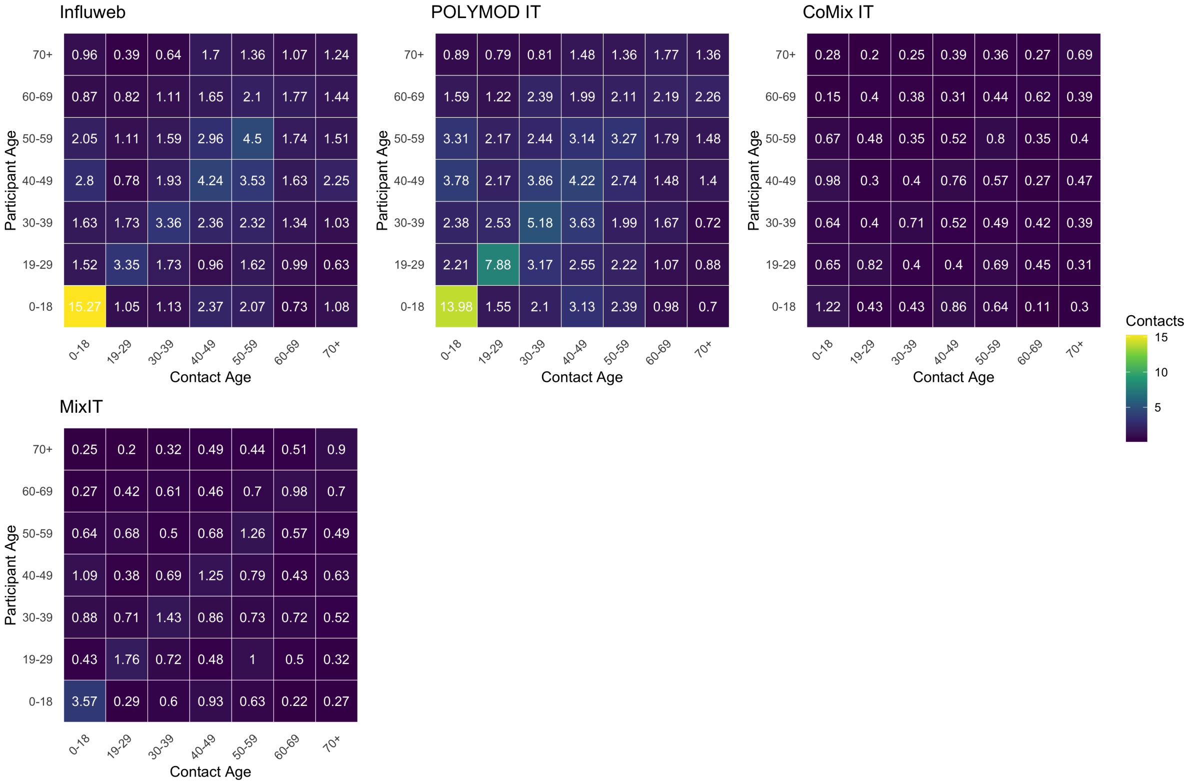


**Fig S1. All-age Italy contact matrices across studies.** Mean bootstrap contact matrices are shown for Influweb, POLYMOD IT, CoMix IT, and MixIT. All panels use the same color scale.

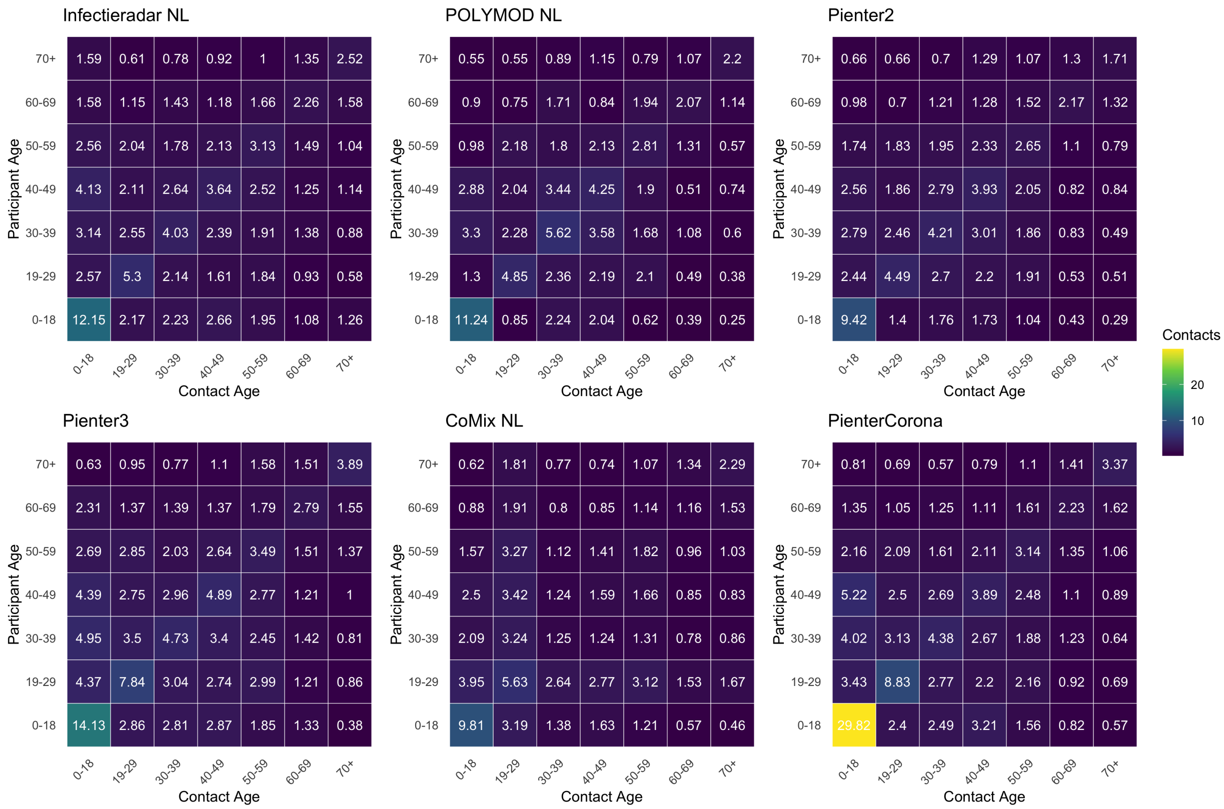


**Fig S2. All-age Netherlands contact matrices across studies.** Mean bootstrap contact matrices are shown for Infectieradar NL, POLYMOD NL, PIENTER 2, PIENTER 3, CoMix NL, and PIENTER Corona. All panels use the same color scale.

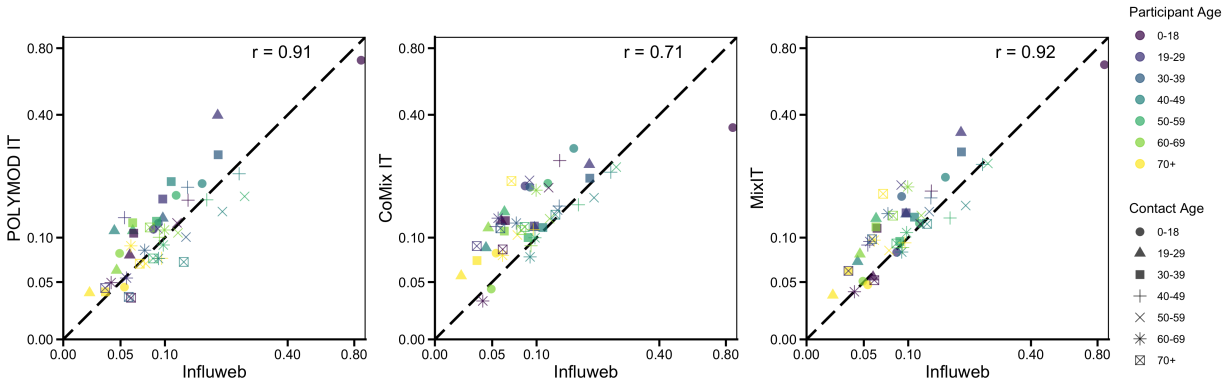


**Fig S3. All-age Italy matrix correlation plots.** Each point represents one matrix cell after dominant-eigenvalue normalization. Influweb is the reference matrix and the dashed line is the identity line.

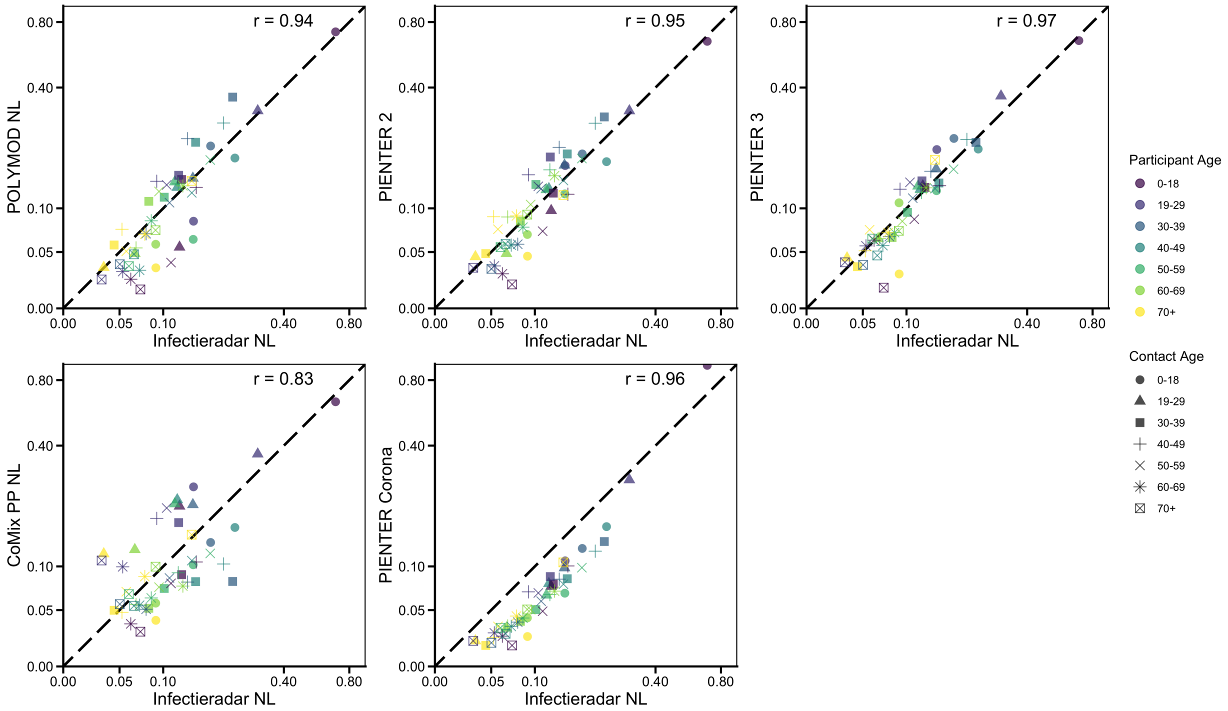


**Fig S4. All-age Netherlands matrix correlation plots.** Each point represents one matrix cell after dominant-eigenvalue normalization. Infectieradar NL is the reference matrix and the dashed line is the identity line.

#### 2.3 Exploring correlates of contact behavior using all ages

We repeated the negative binomial regression models after including participants aged 0–19 years to assess whether excluding minors affected the estimated correlates of contact behavior. We compared the adult-only temporal proximity model in the main text with the all-age temporal proximity model in Table S3, and compared the adult-only symptom burden model in Table S1 with the all-age symptom burden model in Table S4. Overall, including minors did not substantially change the main regression results within countries for either symptom matching approach.

For the temporal proximity models, participants aged 0–19 years had higher contact counts than those aged 50–69 years in Belgium, while most other Belgian coefficients remained similar to the adult-only model. Italy also showed a positive but non-significant coefficient for the 0–19 age group, with otherwise stable estimates; the holiday-period coefficient was somewhat more negative in the all-age model. In the Netherlands, the 0–19 coefficient was positive and marginal, while the other coefficients were very similar to the adult-only model. The negative association for ILI with fever remained strong in Belgium and the Netherlands, and ILI remained associated with a modest reduction in contacts in the Netherlands.

For the symptom burden models, the same overall pattern was observed. The all-age Belgium model showed a positive association for the 0–19 age group, and the remaining Belgian estimates were close to the adult-only model. In Italy, the 0–19 coefficient was positive but not significant, and no symptom category was significantly associated with contact counts. In the Netherlands, the 0–19 coefficient was positive and marginal, symptomatic reports not meeting the ILI definition were associated with higher contact counts, and ILI with fever remained associated with fewer contacts. Taken together, these comparisons suggest that excluding minors did not have a large effect on the estimated associations for the adult covariates or symptom variables.

| Variable | BE Coeff | BE IRR | BE P-value | IT Coeff | IT IRR | IT P-value | NL Coeff | NL IRR | NL P-value |
|---|---|---|---|---|---|---|---|---|---|
| Age group (ref: 50–69) | | | | | | | | | |
| 0–19 | 1.417 | 4.124 | 0.033* | 0.216 | 1.241 | 0.471 | 0.160 | 1.174 | 0.084† |
| 20–49 | -0.137 | 0.872 | 0.243 | -0.231 | 0.794 | 0.043* | -0.066 | 0.936 | 0.001** |
| 70+ | 0.140 | 1.150 | 0.286 | -0.171 | 0.843 | 0.292 | -0.028 | 0.972 | 0.327 |
| Gender (ref: Female) | -0.155 | 0.856 | 0.080† | -0.020 | 0.980 | 0.842 | -0.113 | 0.893 | < 0.001*** |
| Education (ref: HS or less) | 0.432 | 1.540 | < 0.001*** | 0.105 | 1.111 | 0.283 | 0.011 | 1.012 | 0.508 |
| Employment (ref: Employed) | | | | | | | | | |
| Retired | -0.502 | 0.606 | < 0.001*** | -0.683 | 0.505 | < 0.001*** | -0.254 | 0.776 | < 0.001*** |
| Student | -0.398 | 0.672 | 0.477 | -0.107 | 0.898 | 0.597 | 0.077 | 1.080 | 0.294 |
| Unemployed | -0.142 | 0.867 | 0.410 | -0.806 | 0.446 | < 0.001*** | -0.417 | 0.659 | < 0.001*** |
| Household Size | 0.143 | 1.153 | < 0.001*** | -0.009 | 0.991 | 0.687 | 0.141 | 1.151 | < 0.001*** |
| Chronic Condition (ref: False) | 0.130 | 1.139 | 0.203 | 0.124 | 1.132 | 0.234 | -0.055 | 0.946 | 0.002** |
| Holiday period (ref: False) | -0.264 | 0.768 | < 0.001*** | -0.314 | 0.731 | < 0.001*** | -0.099 | 0.905 | < 0.001*** |
| Survey day (ref: Weekend) | 0.065 | 1.068 | 0.451 | 0.293 | 1.341 | < 0.001*** | 0.055 | 1.056 | < 0.001*** |
| Symptoms (ref: None) | | | | | | | | | |
| Symptomatic, no ILI | 0.151 | 1.163 | 0.210 | 0.037 | 1.038 | 0.682 | -0.006 | 0.994 | 0.726 |
| ILI | 0.056 | 1.057 | 0.800 | 0.246 | 1.278 | 0.182 | -0.053 | 0.949 | 0.049* |
| ILI with fever | -1.669 | 0.188 | < 0.001*** | -0.023 | 0.978 | 0.910 | -0.354 | 0.702 | < 0.001*** |

† $p < 0.10$, * $p < 0.05$, ** $p < 0.01$, *** $p < 0.001$

**Table S3. Negative binomial regression results prioritizing temporal proximity of symptoms, all ages.** Results from the negative binomial regression of contact counts from the Infectieradar platform in Belgium, Influweb in Italy, and the Infectieradar platform in the Netherlands, including random intercepts for participants. These models include all ages in the sample. Estimates were obtained using post-stratification weights for age, sex, and day of week. The incidence rate ratio (IRR) is the exponentiated coefficient. The IT model includes a zero-inflation intercept.

| Variable | BE Coeff | BE IRR | BE P-value | IT Coeff | IT IRR | IT P-value | NL Coeff | NL IRR | NL P-value |
|---|---|---|---|---|---|---|---|---|---|
| Age group (ref: 50–69) | | | | | | | | | |
| 0–19 | 1.466 | 4.333 | 0.027* | 0.201 | 1.222 | 0.503 | 0.157 | 1.170 | 0.090† |
| 20–49 | -0.145 | 0.865 | 0.220 | -0.232 | 0.793 | 0.043* | -0.074 | 0.928 | < 0.001*** |
| 70+ | 0.143 | 1.153 | 0.280 | -0.169 | 0.845 | 0.300 | -0.024 | 0.976 | 0.399 |
| Gender (ref: Female) | -0.155 | 0.856 | 0.081† | -0.024 | 0.976 | 0.809 | -0.108 | 0.897 | < 0.001*** |
| Education (ref: HS or less) | 0.435 | 1.545 | < 0.001*** | 0.101 | 1.106 | 0.305 | 0.010 | 1.010 | 0.560 |
| Employment (ref: Employed) | | | | | | | | | |
| Retired | -0.500 | 0.607 | < 0.001*** | -0.690 | 0.502 | < 0.001*** | -0.252 | 0.778 | < 0.001*** |
| Student | -0.423 | 0.655 | 0.451 | -0.109 | 0.896 | 0.590 | 0.068 | 1.070 | 0.356 |
| Unemployed | -0.111 | 0.895 | 0.526 | -0.827 | 0.438 | < 0.001*** | -0.419 | 0.658 | < 0.001*** |
| Household Size | 0.148 | 1.160 | < 0.001*** | -0.008 | 0.992 | 0.746 | 0.140 | 1.151 | < 0.001*** |
| Chronic Condition (ref: False) | 0.143 | 1.154 | 0.164 | 0.123 | 1.131 | 0.237 | -0.059 | 0.943 | 0.001** |
| Holiday period (ref: False) | -0.267 | 0.766 | < 0.001*** | -0.308 | 0.735 | < 0.001*** | -0.099 | 0.906 | < 0.001*** |
| Survey day (ref: Weekend) | 0.076 | 1.079 | 0.384 | 0.294 | 1.341 | < 0.001*** | 0.054 | 1.056 | < 0.001*** |
| Symptoms (ref: None) | | | | | | | | | |
| Symptomatic, no ILI | 0.055 | 1.056 | 0.576 | 0.072 | 1.075 | 0.280 | 0.065 | 1.067 | < 0.001*** |
| ILI | 0.174 | 1.190 | 0.272 | 0.030 | 1.031 | 0.813 | 0.016 | 1.016 | 0.410 |
| ILI with fever | -0.629 | 0.533 | 0.032* | 0.015 | 1.015 | 0.901 | -0.070 | 0.932 | 0.013* |

† $p < 0.10$, * $p < 0.05$, ** $p < 0.01$, *** $p < 0.001$

**Table S4. Negative binomial regression results prioritizing symptom burden, all ages.** Results from the negative binomial regression of contact counts from the Infectieradar platform in Belgium, Influweb in Italy, and the Infectieradar platform in the Netherlands, including random intercepts for participants. These models include all ages in the sample. Estimates were obtained using post-stratification weights for age, sex, and day of week. The incidence rate ratio (IRR) is the exponentiated coefficient. The IT model includes a zero-inflation intercept.

#### 3 Comparison across studies of setting-specific matrices

To provide further context to the comparison across studies, contact matrices stratified by setting were constructed using adult participants, post-stratification weights, and bootstrap sampling. No reciprocity condition was applied. Matrices were then compared using quantitative metrics described in the main text. Matrices constructed for home, work, and leisure settings are shown by country in Figs. S6, S5, and S7, and the corresponding comparison metrics are shown in Table S5.

Across countries, setting-specific comparisons showed the clearest agreement for home contacts. This pattern is expected, as home contacts are structured by household composition and therefore tend to show stable age-mixing patterns across studies. Work contacts also showed broadly comparable structure, although agreement varied more across comparator datasets, likely reflecting differences in employment composition, survey timing, and how work-related contacts were reported. Leisure contacts showed the greatest variability, particularly in overlap-based metrics, suggesting that contacts outside home and work are more sensitive to study context, contact definitions, and time-period-specific social behavior.

Overall, these setting-specific comparisons support the main matrix comparison results that the participatory surveillance platforms capture recognizable age-mixing structure within major contact settings, while differences in absolute intensity were more apparent for settings that are less constrained by household or workplace structure.

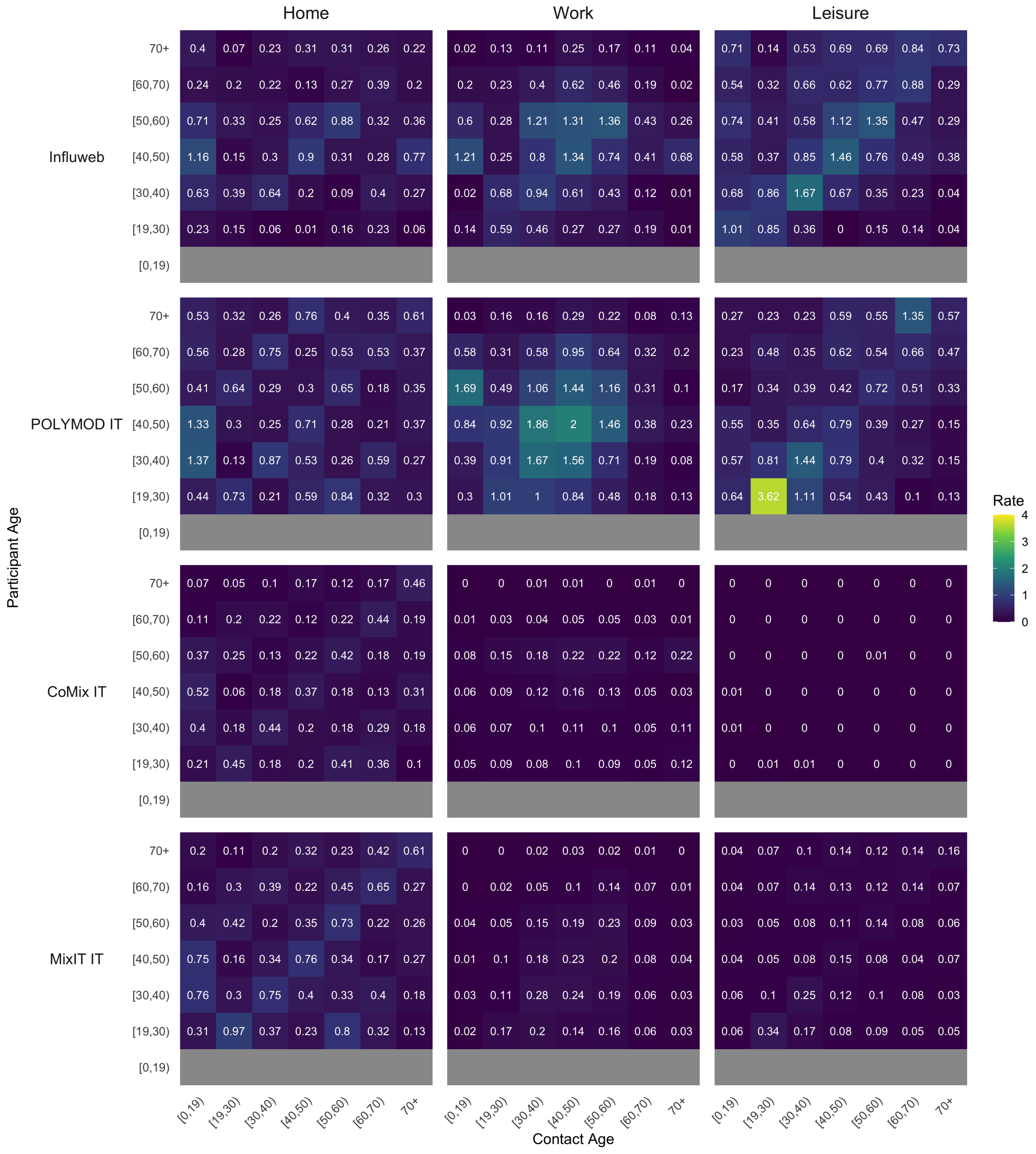


**Fig S5. Italy setting-specific contact matrices for adults across studies.** Mean bootstrapped home, work, and leisure contact matrices are shown for Italian studies included in the setting-specific comparison.

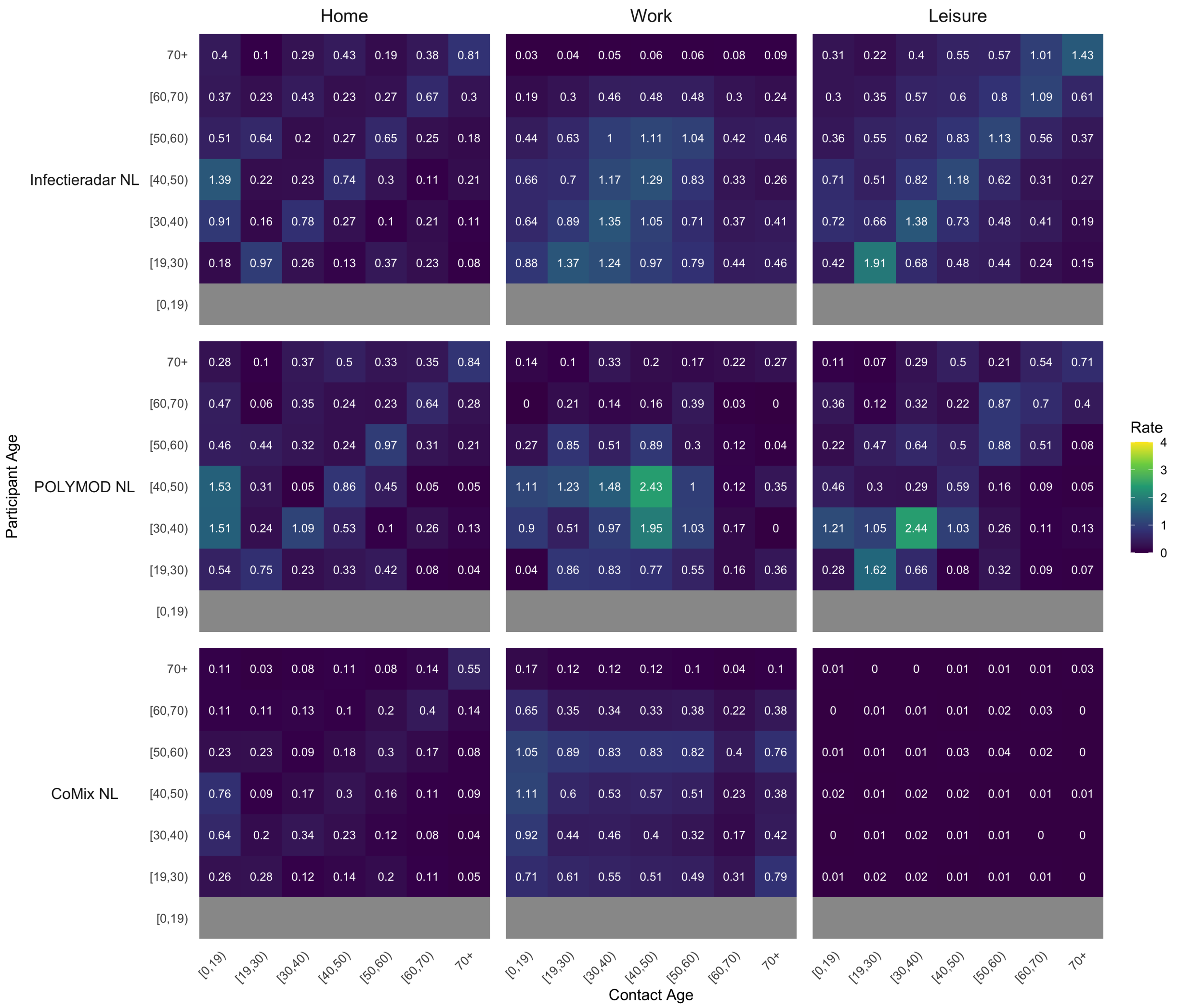


**Fig S6. Netherlands setting-specific contact matrices for adults across studies.** Mean bootstrapped home, work, and leisure contact matrices are shown for Dutch studies included in the setting-specific comparison.

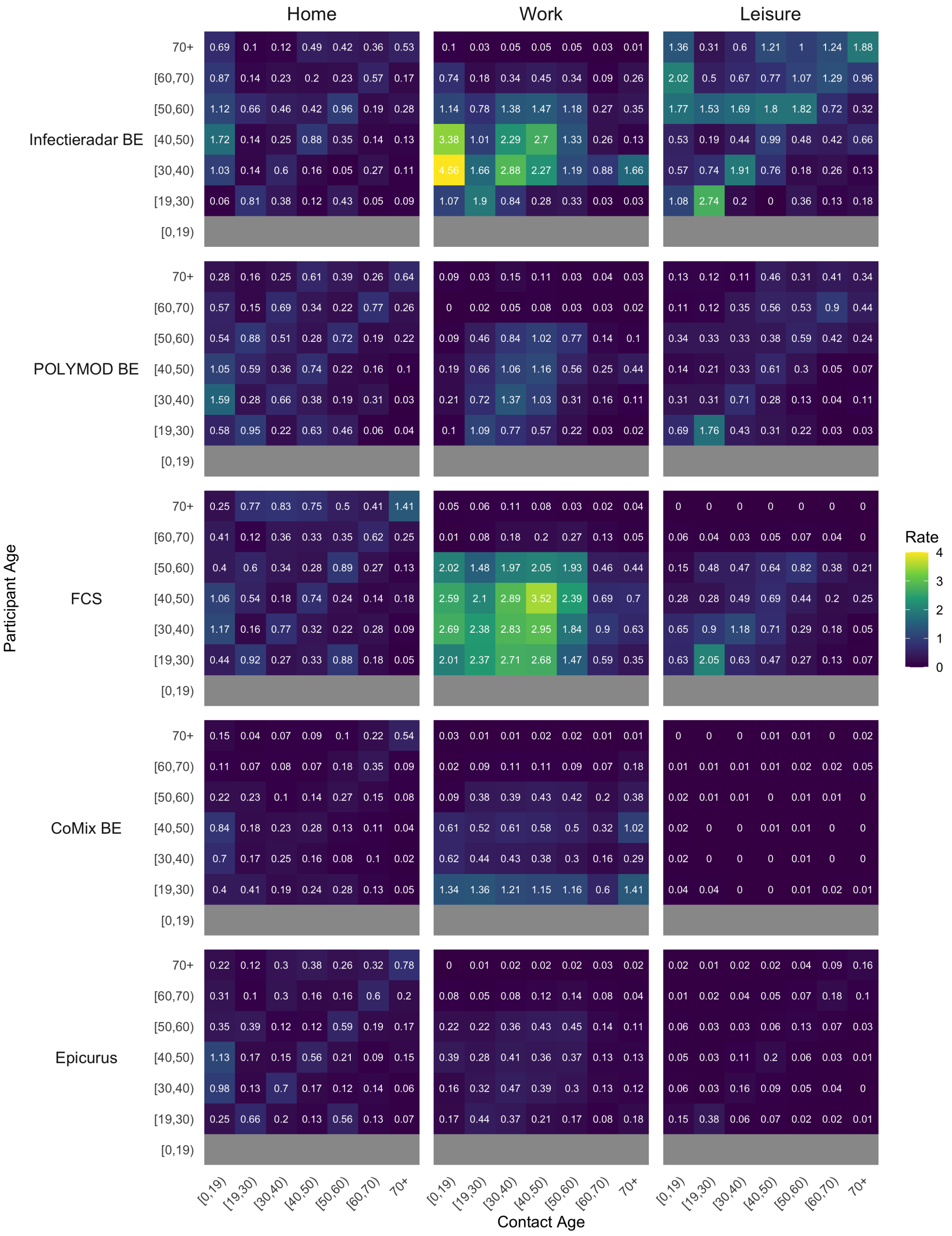


**Fig S7. Belgium setting-specific contact matrices across studies.** Mean bootstrapped home, work, and leisure contact matrices are shown for Belgian studies included in the setting-specific comparison.

### 4 Estimation of bootstrapped $R_0$ distributions across studies

As an additional analysis, we translated bootstrapped contact matrices into scenario-based distributions of the basic reproduction number ($R_0$) under fixed epidemiological assumptions. For each study, $R_0$ was calculated for every bootstrap contact matrix generated during matrix construction. The calculation of the spectral radius requires square matrices, so this analysis was conducted using the adult-by-adult contact matrices.

For each matrix, $R_0$ was calculated as:

$$R_0 = \frac{\beta}{\mu} \cdot \rho(C),$$

where $\rho(C)$ denotes the spectral radius of the unadjusted contact matrix $C$. The transmission and recovery parameters ($\beta = 0.03$ and $\mu = 0.2$) were fixed to represent a respiratory pathogen such as SARS-CoV-2, based on values reported in prior literature [6;7]. We assumed a fully susceptible population at baseline and did not incorporate prior immunity or age-specific susceptibility in order to minimize additional assumptions.

These scenario-based estimates, presented for each country in Figs. S8–S10, were used to illustrate how changes in social contact patterns observed across studies and time periods may translate into differences in transmission potential under a common epidemiological framework. The analysis demonstrates that transmission potential is not static, but varies with the social and epidemiological context captured by each survey. This temporal variability highlights the value of maintaining ongoing participatory surveillance systems, such as the InfluenzaNet platforms, which can provide regularly updated contact data reflecting current population behavior. Because the derived $R_0$ values depend on fixed pathogen assumptions and are influenced by differences in survey design and study context, they should not be interpreted as directly comparable estimates of infectiousness or as absolute measures of transmission potential.

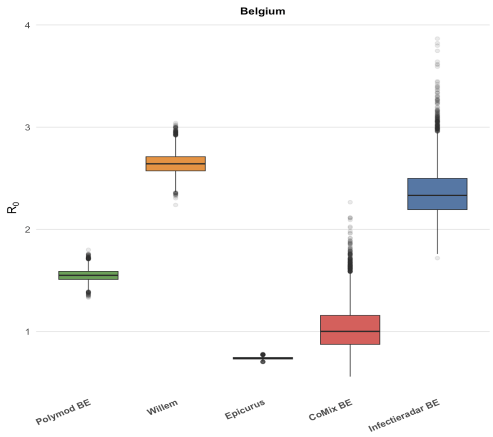


**Fig S8. Belgian bootstrapped $R_0$ distributions across studies.** Distributions of derived $R_0$ values based on Belgian contact matrices from Infectieradar BE and comparator studies.

**Table S5. Setting-specific contact matrix comparison metrics by country.** Metrics compare each setting-specific matrix with the country-specific participatory surveillance reference matrix.

| Setting | Comparator | Cosine similarity | Sørensen–Dice index | Frobenius norm |
|---|---|---|---|---|
| **Belgium** (reference: Infectieradar BE) | | | | |
| *Home* | | | | |
| | CoMix BE | 0.871 | 0.604 | 2.195 |
| | Epicurus BE | 0.918 | 0.779 | 1.534 |
| | POLYMOD BE | 0.887 | 0.784 | 1.664 |
| | FHS BE | 0.843 | 0.754 | 2.008 |
| *Work* | | | | |
| | CoMix BE | 0.606 | 0.441 | 7.440 |
| | Epicurus BE | 0.836 | 0.333 | 7.841 |
| | POLYMOD BE | 0.764 | 0.521 | 6.846 |
| | FHS BE | 0.886 | 0.745 | 5.076 |
| *Leisure* | | | | |
| | CoMix BE | 0.693 | 0.022 | 7.068 |
| | Epicurus BE | 0.804 | 0.138 | 6.642 |
| | POLYMOD BE | 0.852 | 0.539 | 4.862 |
| | FHS BE | 0.710 | 0.498 | 5.297 |
| **Italy** (reference: Influweb) | | | | |
| *Home* | | | | |
| | CoMix IT | 0.876 | 0.702 | 1.476 |
| | MixIT IT | 0.845 | 0.763 | 1.559 |
| | POLYMOD IT | 0.849 | 0.713 | 1.920 |
| *Work* | | | | |
| | CoMix IT | 0.843 | 0.278 | 3.296 |
| | MixIT IT | 0.856 | 0.337 | 3.172 |
| | POLYMOD IT | 0.906 | 0.731 | 2.684 |
| *Leisure* | | | | |
| | CoMix IT | 0.776 | 0.007 | 4.561 |
| | MixIT IT | 0.870 | 0.273 | 3.946 |
| | POLYMOD IT | 0.759 | 0.742 | 3.440 |
| **Netherlands** (reference: Infectieradar NL) | | | | |
| *Home* | | | | |
| | CoMix NL | 0.952 | 0.664 | 1.597 |
| | POLYMOD NL | 0.957 | 0.849 | 1.080 |
| *Work* | | | | |
| | CoMix NL | 0.866 | 0.743 | 2.340 |
| | POLYMOD NL | 0.857 | 0.714 | 2.554 |
| *Leisure* | | | | |
| | CoMix NL | 0.911 | 0.037 | 4.639 |
| | POLYMOD NL | 0.886 | 0.757 | 2.194 |

Higher cosine similarity and Sørensen–Dice index indicate greater similarity; lower Frobenius norm indicates smaller absolute matrix differences. Metrics are rounded to three decimal places.

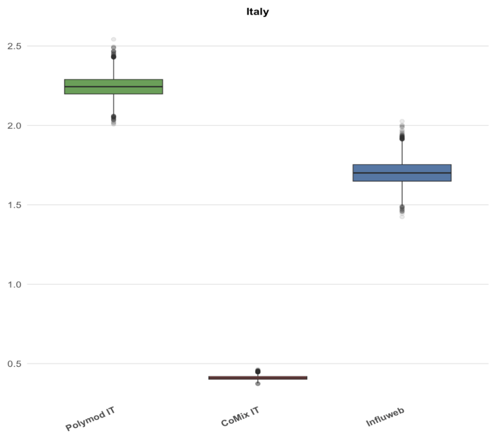


**Fig S9. Italian bootstrapped $R_0$ distributions across studies.** Distributions of derived $R_0$ values based on Italian contact matrices from Influweb and comparator studies.

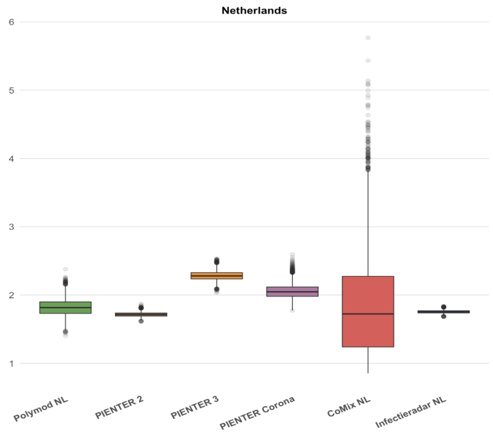


**Fig S10. Dutch bootstrapped $R_0$ distributions across studies.** Distributions of derived $R_0$ values based on Dutch contact matrices from Infectieradar NL and comparator studies.
